# Waist, waist-height-ratio vs body mass index and the risks of multiple diseases: a cohort study with replication

**DOI:** 10.1101/2025.10.16.25338152

**Authors:** Frederick Ho, Jaana Pentti, Mika Kivimäki, Naveed Sattar

## Abstract

**Importance:** Body mass index (BMI) is widely used to assess obesity-related health risks, but waist circumference (WC) and waist-to-height ratio (WHtR) are thought to better capture central adiposity and associated disease risk.

**Objective:** To compare the associations of BMI, WC, and WHtR with the risk of 78 incident conditions, multiple long-term conditions (MLTCs), and mortality, and to evaluate whether waist-based measures improve risk identification over BMI and, if so, over which BMI range.

**Design, Setting, and Participants:** This cohort study analyzed data from 495,911 adults in the UK Biobank with a mean follow-up of 13.7 years. Findings were replicated in the Whitehall II study (n = 7,973; mean follow-up, 28.6 years).

**Exposures:** Obesity defined by BMI, WC, and WHtR.

**Main Outcomes and Measures:** Associations of each adiposity measure with 78 incident conditions, MLTCs, and mortality were assessed using Cox proportional hazards models. Population attributable fractions (PAFs) were calculated to estimate the proportion of outcomes attributable to obesity defined by each measure.

**Results:** Obesity defined by BMI, WC, or WHtR was associated with increased risk for most outcomes. WHtR-defined obesity generally showed the strongest associations, followed by WC and BMI, with some exceptions (e.g., osteoarthritis). However, PAFs for MLTCs for WC and WHtR were <1% percentage points higher than for BMI. Notably, meaningful gains in PAFs were observed <u>only</u> among individuals with BMI between 25.0 and 29.9. These findings were replicated in the Whitehall II cohort.

**Conclusions and Relevance:** Replacing BMI with waist-based measures **did not** meaningfully improve risk identification for MLTCs in the general population when BMI was ≥30. However, waist-based measures may offer additional predictive value among individuals with BMI <30.

**Key Points:** *Question:* Do waist-based measures (waist circumference [WC] and waist-to-height ratio [WHtR]) explain the risk of multiple long-term conditions (MLTCs) and mortality better than body mass index (BMI), and if so, in which BMI ranges?

*Findings:* In two cohort studies of nearly 500,000 UK adults, WHtR and WC were generally more strongly associated with incident conditions, MLTCs, and mortality than BMI. However, the improvement was modest overall and primarily evident in individuals with BMI between 25.0 and 29.9.

*Meaning:* Waist-based measures may have a stronger association over BMI in individuals with overweight (BMI <30), but the difference was marginal and non-significant in those with a BMI ≥30.

## Introduction

A recent study reported associations between BMI and incident outcomes across many diseases, demonstrating that obesity is strongly linked to the risk for developing multiple long-term conditions (MLTCs).^1^ The population attributable fraction (PAF) for BMI ≥30 compared to normal BMI was 55% for MLTCs, suggesting obesity as the most important modifiable risk factor, a finding with major implications for health policy.

At the same time, the merits of BMI as a predictor of health risk continues to be debated, with many advocating the much wider use of other simple adiposity measures, such as waist circumference (WC) and waist to height ratio (WHtR), that better capture adiposity distribution and may more accurately predict obesity-related adverse outcomes,^2,3^ including the onset of MLTCs. A recent Lancet commission also recommended that clinical assessment of obesity should shift more towards using WC or WHtR, at least up to BMI levels ≥40, whereas others (e.g. NICE) suggest the relevant threshold should be ≥35 above which BMI alone suffices to categorise obesity. However, these considerations and central measures of obesity are not yet standard practice in most clinical settings.^4^

The present study compared the associations between each of these measures and obesity-related outcomes, using z-scores to directly compare their relative performance. It examined how well each of the three adiposity measures predicted incident MLTCs and to what extent measurements of WC or WHtR improved PAF for obesity-related MTLCs beyond that explained by BMI alone. In addition, the analyses were repeated by subgrouping individuals into specific BMI categories of clinical relevance to agnostically examine below which BMI category measures of centrally obesity might add prognostic value for MLTCs. The hypothesis was this would be close to a BMI of 35, given NICE suggests no need to consider other obesity measures above this cut-off. The findings were validated in a separate smaller cohort but with much longer follow-up.

## Methods

### Study population

The main analyses were based on the UK Biobank study, with replication analyses conducted in the Whitehall study. Over 500,000 participants aged 37-73 years participated in the UK Biobank prospective cohort study (5.5% response rate). They attended 1 of 22 assessment centres across England, Wales, and Scotland between 2006 and 2010. All participants completed touchscreen questionnaires and provided baseline physical measurements and biological samples. The UK Biobank study was approved by the North West Multi-Centre Research Ethics Committee (Ref 11/NW/0382 on June 17, 2011). All participants provided written informed consent to participate. The study protocol is available online (http://www.ukbiobank.ac.uk/). The present research uses the UK Biobank resource under application number 71392.

The Whitehall II study is a prospective occupational cohort study involving 10,308 London-based civil servants aged 35–55, with baseline data collected between 1985 and 1988 (response rate: 74.0%). Measurements of BMI, WC, and WHtR were first obtained during the 1991–1994 clinical examination, which included 8,815 participants (86.6% of those still alive from the original cohort). All participants were linked to NHS electronic health records. Written informed consent was obtained from all participants, and ethical approval was granted by the University College London Hospital Committee on the Ethics of Human Research (reference: 85/0938).

We excluded participants with missing weight, WC, hip circumference (HC), or height in all analyses and people who had prior diseases or MLTCs for disease and MLTC incidence analysis.

### Outcome: 78 diseases, multimorbidity and all-cause mortality

In both studies, participants were linked to the UK National Health Service (NHS) Hospital Episode Statistics (HES) database for hospital admissions and the NHS Central Registry for mortality. Similar to a previous study,^1^ diseases were coded based on WHO’s International Classification of Diseases 10th Revision (ICD-10)(**Supplement**). We focused on a predefined list of 78 common ICD-10 disease chapters and diagnostic groups constructed for outcome-wide studies by investigators who were masked to exposure data (including BMI). Three definitions of multimorbidity were included: simple MLTC (incidence of 2 conditions); incidence of 3 conditions; and complex MLTC (incidence of 4 conditions), all based on 21 obesity-related conditions.^1^

### Exposures: BMI, WC, and WHtR

Body weight, WC, and height were measured by trained staff using standard methods and equipment. BMI was calculated from weight (in kilograms) divided by the square of height (in meters).^5^ Waist-to-height ratio (WHtR) was calculated from waist circumference divided by height. These were used as both numeric and categorical variables. The WHO definition of overweight (BMI≥25 and <30) and obesity (BMI≥30) was used. Sex-specific WC and WHtR cutpoints were chosen so that WC/WHtR-defined overweight and obesity have similar prevalence as BMI-defined ones. In the UK Biobank, the cutpoints for WC were 93 cm for female and 103 cm for male, and those for WHtR were 0.57 for female and 0.59 for male. The corresponding values in the Whitehall study were 88 cm and 103 cm for WC and 0.55 and 0.58 for WHtR. The International Diabetes Foundation cutpoints and externally derived cutpoints based on the population representative HES-SHS (2005-2019) were used as a sensitivity analysis. All cutpoints are shown in Supplement eTable 1.

### Baseline covariates

Age at baseline was calculated from date of birth and date of baseline assessment. Highest level of education (UK Biobank) and employment grade (Whitehall), ethnicity and sex were self-reported. Ethnicity was categorised as: White, South Asian, Black, Chinese, mixed and other in the UK Biobank and, due to smaller sample size, White vs non-White in the Whitehall study. In both studies, the Townsend deprivation index, an area-based measure of socioeconomic status, was derived from postcode of residence using aggregated data on unemployment, car and home ownership and household overcrowding.^6^ Alcohol intake was self-reported as units of consumption per week. Smoking status was self-reported at baseline as never, previous and current. Physical activity was calculated from the frequency and duration of walking, moderate and vigorous activity, reported using the international physical activity questionnaire, and converted into total metabolic equivalent of task (MET) per week.^7^ Dietary quality was measured using a 9-point score based on national dietary guidelines.^8^ Additional details about these measurements can be found in the UK Biobank online protocol.

### Statistical analyses

The characteristics of the cohort were summarised as mean and standard deviation (SD) for continuous variables and frequency and percentage (%) for categorical variables. Nonlinear associations of adiposity with incident diseases were investigated using Cox proportional hazard models with penalised splines.^9^ Adiposity variables were standardised as sex-specific z-score (mean=0; SD=1) so that they are on the same scale for comparison. Z-scores are zeroed at 27.2 kg/m^2^ for female and 27.9 kg/m^2^ for male in BMI; 83.9 cm for female and 97.0 cm for male in WC; and 0.52 for female and 0.55 for male in WHtR. One z-score unit is equivalent to 5.15 kg/m^2^ for female and 4.22 kg/m^2^ for male in BMI; 12.5 cm for female and 11.3 cm for male in WC; 0.079 for female and 0.065 for male in WHtR. Likelihood ratio tests were used to compare the model’s using splines and those assuming linearity. The primary analyses were adjusted for potential confounding factors, including age, sex, ethnicity, education/employment status, deprivation, physical activity, dietary quality score, smoking status, and alcohol intake. Similarly categorical variables of BMI, WC, and WHtR were analysed. To avoid over adjustment such analyses were also repeated with only age, sex and ethnicity adjustments as supplementary.

PAF based on the hazard ratio (HR) of the categorical analyses were estimated. The differences in PAFs between WC and BMI and between WHtR and BMI were used to estimate to extent to which additional cases that could be identified/prevented due to the change from BMI-based obesity (BMI≥30) to other equivalent measures. Additionally, the same analysis was replicated for overweight (BMI≥25 and <30), class I obesity (BMI≥30 and <35) and class 2 and 3 obesity (BMI≥35), comparing with the equivalents. Lastly, as a sensitivity analysis, the results of the primary outcomes (mortality and multimorbidity) were repeated with adjustment of only age, sex, and ethnicity.

Statistical analyses were performed using the statistical software R 4.2.2 with the survival packages in the UK Biobank and SAS (9.4) in the Whitehall study. Holm’s Bonferroni procedure was used to correct p-values for multiple testing.^10^

## Results

After excluding UK Biobank participants with missing data, 495,911 were included in the analysis. Participant characteristics by WC-defined obesity categories are shown in **Table 1** (part A). Those with WC-defined obesity were slightly older, more likely to be male, more deprived, less physically active, had lower diet quality, were less likely to be never smokers, and consumed more alcohol.

**Table 1.** Participant characteristics at baseline.

|  | WC categories based on equivalent BMI |  |  |
| --- | --- | --- | --- |
|  | Normal weight<br>equivalent | Overweight<br>equivalent | Obesity<br>equivalent |
| **UK Biobank** |  |  |  |
| N | 152,769 | 214,783 | 128,359 |
| Mean (SD) age (years) | 55 (8) | 57 (8) | 58 (8) |
| Sex |  |  |  |
| Female | 102,298 (67%) | 101,274 (47%) | 65,799 (51%) |
| Male | 50,471 (33%) | 113,509 (53%) | 62,560 (49%) |
| Ethnicity |  |  |  |
| White | 144,576 (95%) | 202,237 (95%) | 120,527 (94%) |
| Non-White | 7588 (5%) | 11,525 (5%) | 7126 (6%) |
| Education |  |  |  |
| University/College | 58,905 (39%) | 68,266 (32%) | 31,881 (25%) |
| Others | 92,758 (61%) | 144,690 (68%) | 95,213 (75%) |
| Mean (SD) deprivation<br>index | -1.52 (3.00) | -1.42 (3.02) | -0.86 (3.25) |
| Mean (SD) physical activity<br>(MET-minutes/week) | 2,634 (2,450) | 2,403 (2,412) | 1,986 (2,250) |
| Mean (SD) diet quality | 4.59 (1.53) | 4.32 (1.50) | 4.13 (1.48) |
| Smoking status |  |  |  |
| Never | 93,250 (61%) | 116,279 (54%) | 63,131 (49%) |
| Previous | 43,666 (29%) | 76,027 (35%) | 51,747 (40%) |
| Current | 15,853 (10%) | 22,477 (10%) | 13,481 (11%) |
| Mean (SD) alcohol<br>consumptions (units/week) | 14 (15) | 17 (19) | 16 (20) |
| **Whitehall** |  |  |  |
| N | 4154 | 3037 | 782 |
| Mean (SD) age (years) | 49 (6) | 51 (6) | 52 (6) |
| Sex |  |  |  |
| Female | 1264 (30) | 808 (27) | 391 (50) |
| Male | 2890 (70) | 2229 (73) | 391 (50) |
| Ethnicity |  |  |  |
| White | 3806 (92) | 2714 (89) | 665 (85) |
| Non-White | 334 (8) | 311 (10) | 115 (15) |
| Missing | 14 (0.3) | 12 (0.4) | 2 (0.3) |
| Employment status |  |  |  |
| High | 1648 (40) | 1117 (37) | 207 (26) |
| Intermediate | 1875 (45) | 1345 (44) | 315 (40) |
| Low | 571 (14) | 495 (16) | 227 (29) |
| Missing | 60 (1) | 80 (3) | 33 (4) |
| Deprivation index |  |  |  |
| Low | 1618 (39) | 1149 (38) | 244 (31) |
| Intermediate | 1384 (33) | 954 (31) | 225 (29) |
| High | 974 (23) | 777 (26) | 280 (36) |
| Missing | 178 (4) | 157 (5) | 33 (4) |
|  | Normal weight<br>equivalent | Overweight<br>equivalent | Obesity<br>equivalent |
| Physical activity |  |  |  |
| Low | 727 (18) | 632 (21) | 247 (32) |
| Intermediate | 1459 (35) | 1067 (35) | 246 (31) |
| High | 1910 (46) | 1260 (41) | 258 (33) |
| Missing | 58 (1) | 78 (3) | 31 (4) |
| Dietary quality |  |  |  |
| Low | 872 (21) | 745 (25) | 192 (25) |
| Intermediate | 2309 (56) | 1646 (54) | 397 (51) |
| High | 912 (22) | 567 (19) | 161 (21) |
| Missing | 61 (1) | 79 (3) | 32 (4) |
| Current smoking |  |  |  |
| No | 3545 (85) | 2565 (84) | 637 (81) |
| Yes | 550 (13) | 393 (13) | 113 (14) |
| Missing | 59 (1) | 79 (3) | 32 (4) |
| Alcohol consumptions<br>(units/week) |  |  |  |
| Non | 767 (18) | 527 (17) | 216 (28) |
| Moderate | 2779 (67) | 1874 (62) | 423 (54) |
| Heavy | 549 (13) | 553 (18) | 111 (14) |
| Missing | 59 (1) | 83 (3) | 32 (4) |

Overweight equivalent: WC $\geq 80$ & $< 93$ (Female); WC $\geq 89$ & $< 103$ (Male) in UK Biobank and WC $\geq 75$ & $< 88$ (Female); WC $\geq 88$ & $< 103$ (Male) in the Whitehall study.
Obesity equivalent: WC $\geq 93$ (Female); WC $\geq 103$ (Male) in UK Biobank and WC $\geq 88$ (Female); WC $\geq 103$ (Male) in the Whitehall study.

During a mean follow-up of 13.7 years (SD 1.96), obesity defined by BMI, WC, or WHtR was associated with higher risks of most outcomes, except prostate cancer, melanoma, and road accidents (**Supplement eTable 2**). WHtR-defined obesity generally showed the strongest associations, followed by WC and BMI (**Table 2**). For example, HRs for diabetes mellitus were 8.87, 7.87, and 6.79 for WHtR-, WC-, and BMI-defined obesity, respectively. Osteoarthritis showed stronger associations with BMI-defined obesity. Similar findings were observed using alternative WC and WHtR cutpoints (**Supplement eTable 3**).

**Table 2.** Association of obesity defined using BMI, WC, and WHtR with mortality, multimorbidity and top 15 obesity-related diseases in the UK Biobank and Whitehall studies.

| Study and outcome | HR for obesity vs normal weight defined using |  |  |
| --- | --- | --- | --- |
|  | BMI | WC | WHtR |
| **UK Biobank** |  |  |  |
| All-cause mortality | 1.31 (1.28-1.35) | 1.42 (1.38-1.46) | 1.47 (1.43-1.51) |
| Simple multimorbidity (2<sup>nd</sup> disease) | 2.38 (2.35-2.41) | 2.44 (2.41-2.48) | 2.49 (2.45-2.52) |
| Third disease | 2.78 (2.73-2.83) | 2.93 (2.88-2.98) | 3.01 (2.96-3.07) |
| Complex multimorbidity (4<sup>th</sup> disease) | 3.19 (3.13-3.26) | 3.45 (3.38-3.53) | 3.60 (3.52-3.68) |
| Top 15 obesity-related diseases |  |  |  |
| Diabetes mellitus | 6.79 (6.56-7.03) | 7.87 (7.58-8.18) | 8.87 (8.53-9.22) |
| Sleep disorders | 6.25 (5.85-6.67) | 6.38 (5.95-6.84) | 6.47 (6.05-6.92) |
| Gout | 4.60 (4.31-4.92) | 4.57 (4.25-4.92) | 4.65 (4.33-4.99) |
| Hypertension | 2.79 (2.75-2.84) | 2.86 (2.81-2.90) | 3.03 (2.98-3.08) |
| Liver diseases | 2.53 (2.42-2.64) | 2.86 (2.73-3.00) | 3.04 (2.90-3.19) |
| Heart failure | 2.45 (2.35-2.55) | 2.57 (2.46-2.69) | 2.57 (2.46-2.68) |
| Osteoarthritis | 2.40 (2.35-2.44) | 2.22 (2.18-2.27) | 2.20 (2.16-2.25) |
| Renal failure | 2.35 (2.28-2.41) | 2.46 (2.39-2.53) | 2.53 (2.46-2.60) |
| Pancreatitis | 2.23 (2.02-2.46) | 2.62 (2.35-2.91) | 2.69 (2.42-2.99) |
| Pulmonary embolism | 2.19 (2.06-2.32) | 2.36 (2.21-2.51) | 2.08 (1.96-2.21) |
| Eczema and skin infections | 2.05 (1.98-2.12) | 2.18 (2.10-2.26) | 2.16 (2.09-2.24) |
| Ischaemic heart disease | 2.00 (1.95-2.05) | 2.03 (1.98-2.08) | 2.18 (2.12-2.24) |
| Deep vein thrombosis | 1.90 (1.79-2.02) | 1.98 (1.86-2.12) | 1.79 (1.68-1.91) |
| Sciatica | 1.86 (1.78-1.94) | 1.80 (1.72-1.88) | 1.78 (1.71-1.87) |
| Kidney cancer | 1.85 (1.64-2.08) | 1.95 (1.72-2.21) | 1.91 (1.69-2.16) |
| **Whitehall** |  |  |  |
| All-cause mortality | 1.73 (1.53-1.96) | 1.70 (1.50-1.92) | 1.77 (1.56-2.00) |
| Simple multimorbidity (2<sup>nd</sup> disease) | 2.41 (2.19-2.66) | 2.50 (2.27-2.76) | 2.47 (2.24-2.73) |
| Third disease | 2.57 (2.30-2.88) | 2.65 (2.37-2.97) | 2.60 (2.32-2.91) |
| Complex multimorbidity (4<sup>th</sup> disease) | 3.21 (2.81-3.66) | 3.21 (2.81-3.66) | 3.18 (2.78-3.63) |
| Top 15 obesity-related diseases |  |  |  |
| Diabetes mellitus | 4.54 (3.86-5.35) | 4.67 (3.96-5.51) | 5.46 (4.61-6.45) |
| Sleep disorders | 4.67 (3.16-6.90) | 5.28 (3.57-7.81) | 4.56 (3.07-6.79) |
| Gout | 5.95 (4.18-8.48) | 4.96 (3.44-7.14) | 5.02 (3.49-7.22) |
| Hypertension | 2.40 (2.17-2.66) | 2.30 (2.08-2.55) | 2.45 (2.21-2.72) |
| Liver diseases | 1.63 (1.18-2.25) | 2.22 (1.65-3.00) | 1.95 (1.43-2.65) |
| Heart failure | 2.71 (2.20-3.33) | 2.84 (2.31-3.49) | 2.73 (2.22-3.35) |
| Osteoarthritis | 2.14 (1.84-2.49) | 2.04 (1.75-2.38) | 2.00 (1.71-2.33) |
| Renal failure | 2.16 (1.83-2.53) | 2.21 (1.88-2.60) | 2.16 (1.83-2.55) |
| Pancreatitis | 2.86 (1.41-5.80) | 3.47 (1.77-6.80) | 2.82 (1.42-5.60) |
| Pulmonary embolism | 2.29 (1.63-3.24) | 2.63 (1.83-3.78) | 1.92 (1.31-2.81) |
| Eczema and skin infections | 1.90 (1.52-2.37) | 1.97 (1.58-2.46) | 1.89 (1.52-2.36) |
| Ischaemic heart disease | 1.90 (1.62-2.21) | 1.97 (1.69-2.29) | 2.09 (1.79-2.42) |
| Deep vein thrombosis | 2.46 (1.71-3.53) | 2.83 (1.96-4.08) | 2.65 (1.85-3.80) |
| Sciatica | 1.21 (0.83-1.77) | 0.98 (0.65-1.47) | 1.07 (0.72-1.59) |
| Kidney cancer | 1.25 (0.58-2.72) | 1.12 (0.49-2.55) | 1.03 (0.42-2.50) |

Numbers presented are HR (95% CI) compared with the normal weight group and its WC/WHtR equivalent. Adjusted for age, sex, ethnicity, education/employment grade, deprivation, physical activity, dietary quality, smoking, and alcohol consumption.

**Figure 1** shows non-linear associations of z-scores of BMI, WC, and WHtR with mortality and MLTCs. WC and WHtR had stronger associations. Notably, BMI z-score < −1 (equivalent to BMI 22 in females, 23 in males) was linked to higher mortality, a pattern not seen for WC or WHtR.

**Figure 1.**
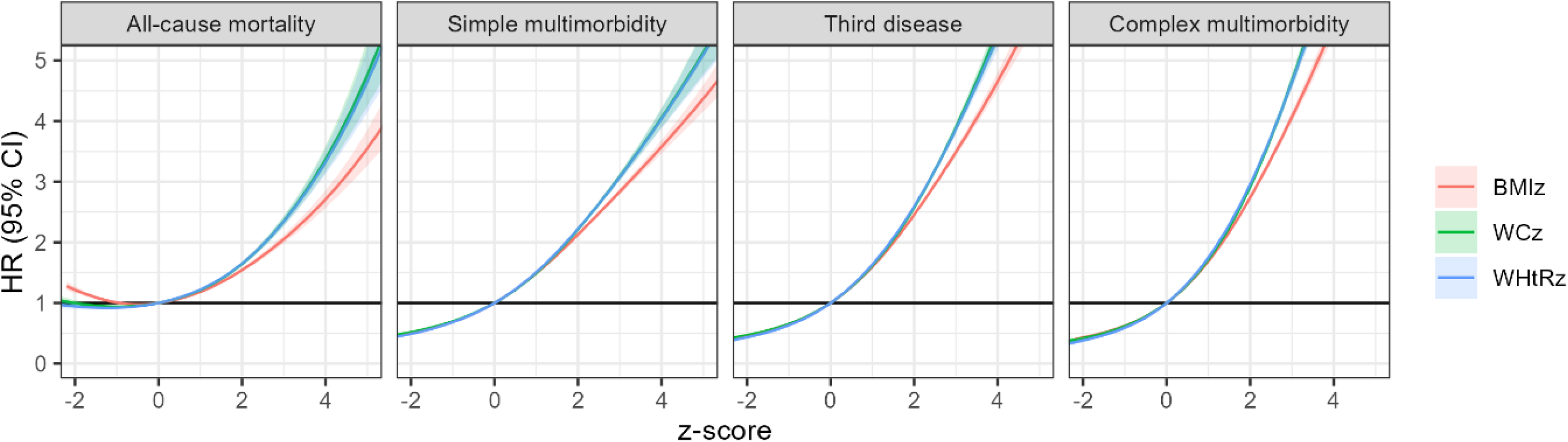
Associations of BMI, WC, and WHtR z-scores with mortality and incident multimorbidity in the UK Biobank. Adjusted for age, sex, ethnicity, education, deprivation, physical activity, dietary quality, smoking, and alcohol consumption. The thresholds for obesity (BMI≥30) are 0.54 for female and 0.50 for male.

**Figure 2** shows non-linear associations with the top 15 obesity-related diseases. WC and WHtR z-scores showed stronger associations than BMI for diabetes, gout, heart failure, hypertension, renal failure, liver disease, eczema and skin infections, ischaemic heart disease, and arrhythmias. Full results are in **Supplement e**Figures 1–7. Log-linear models (**Supplement eTable 4**) showed similar patterns.

**Figure 2.**
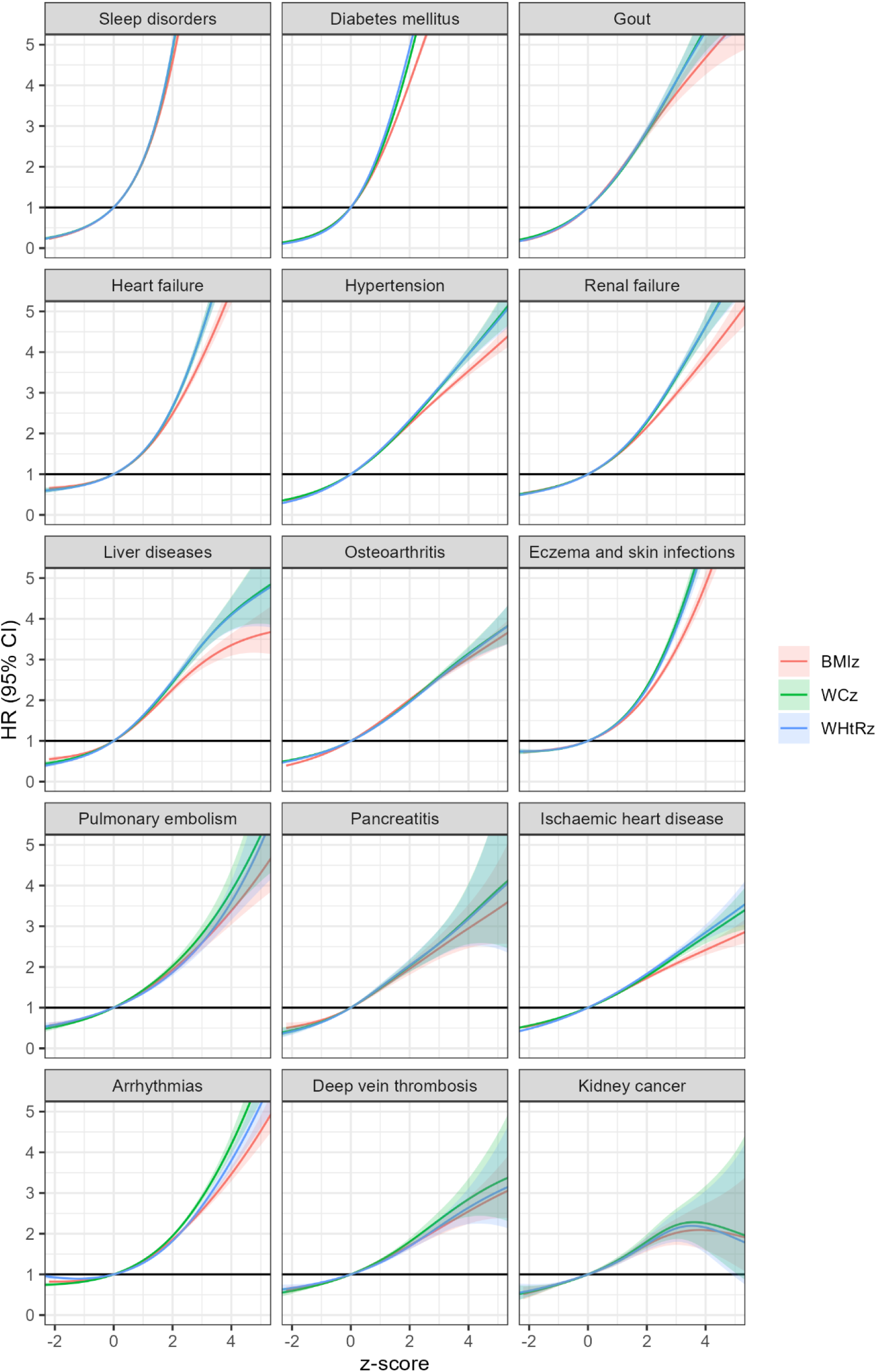
Association of BMI, WC, and WHtR with incidence of top 15 obesity-related diseases in the UK Biobank. Adjusted for age, sex, ethnicity, education, deprivation, physical activity, dietary quality, smoking, and alcohol consumption

For all-cause mortality, WC- and WHtR-defined obesity had only marginally higher PAFs than BMI, with differences of 0.18 and 0.19 percentage points, respectively, in those with BMI ≥30 (**Table 3**). PAF differences for MLTCs were similarly minimal, all <1% and not statistically significant (**Supplement eTable 5**). Similar findings were seen across obesity classes (Supplement eTable 6).

**Table 3.** Differences in population attributable fraction (PAF) for mortality and multimorbidity between obesity and overweight defined using WC, WHtR, and BMI.

| Study and outcome | Obesity (BMI ≥ 30) |  |  |  |  |  | Overweight (BMI ≥ 25 & < 30) |  |  |  |  |  |
| --- | --- | --- | --- | --- | --- | --- | --- | --- | --- | --- | --- | --- |
|  | PAF<sub>WC</sub> - PAF<sub>BMI</sub> |  |  | PAF<sub>WHtR</sub> - PAF<sub>BMI</sub> |  |  | PAF<sub>WC</sub> - PAF<sub>BMI</sub> |  |  | PAF<sub>WHtR</sub> - PAF<sub>BMI</sub> |  |  |
|  | % | Lower 95% CI | Upper 95% CI | % | Lower 95% CI | Upper 95% CI | % | Lower 95% CI | Upper 95% CI | % | Lower 95% CI | Upper 95% CI |
| **UK Biobank** |  |  |  |  |  |  |  |  |  |  |  |  |
| All-cause mortality | 0.18 | -1.97 | 2.34 | 0.19 | -1.97 | 2.35 | 5.35 | 3.28 | 7.42 | 7.17 | 5.13 | 9.22 |
| Simple multimorbidity (2nd disease) | 0.06 | -0.76 | 0.87 | 0.07 | -0.75 | 0.88 | 2.13 | 1.27 | 3.00 | 2.84 | 1.99 | 3.69 |
| Third disease | 0.14 | -0.85 | 1.13 | 0.15 | -0.84 | 1.14 | 4.09 | 2.91 | 5.28 | 5.04 | 3.87 | 6.21 |
| Complex multimorbidity (4th disease) | 0.17 | -1.00 | 1.35 | 0.19 | -0.99 | 1.37 | 5.76 | 4.13 | 7.38 | 7.16 | 5.56 | 8.77 |
| **Whitehall** |  |  |  |  |  |  |  |  |  |  |  |  |
| All-cause mortality | -0.56 | -1.85 | 0.60 | 0.18 | -1.10 | 1.38 | 3.95 | 1.20 | 7.19 | 1.76 | -1.09 | 4.91 |
| Simple multimorbidity (2nd disease) | 0.31 | -1.01 | 1.77 | 0.35 | -1.11 | 1.84 | 1.93 | -0.33 | 3.83 | 0.53 | -1.54 | 2.77 |
| Third disease | 0.22 | -1.60 | 1.68 | 0.05 | -1.48 | 1.55 | 1.96 | -0.88 | 4.71 | 1.19 | -1.37 | 3.65 |
| Complex multimorbidity (4th disease) | -0.35 | -2.54 | 1.33 | -0.39 | -2.39 | 1.36 | 1.89 | -1.71 | 4.92 | 1.68 | -1.49 | 4.96 |

PAF assumes the HR estimated to be causal which may not be true.
HRs were adjusted for age, sex, ethnicity, education/employment grade, deprivation, physical activity, dietary quality, smoking, and alcohol consumption.

However, WC- and WHtR-defined overweight showed notably higher PAFs than BMI-defined overweight. WHtR-defined overweight had >7% higher PAFs for all-cause mortality and complex multimorbidity (**Supplement eTable 7, eTable 3**).

The replication analysis included 7,973 Whitehall participants with complete obesity data and a mean follow-up of 28.6 years (SD 6.0). Compared to UK Biobank, Whitehall participants were younger (mean age 50.1 vs 55 years), more often male (69.1% vs 45.7%), and less likely to have obesity by BMI (9.8% vs 25.9%) (**Table 1, part** B).

Obesity defined by BMI, WC, and WHtR showed similarly strong associations with all-cause mortality (HRs: 1.73, 1.70, and 1.77, respectively). HRs for MLTCs and top 15 obesity-related diseases were also similar across definitions: 2.41–2.50 for simple MLTCs, 2.57–2.65 for the third disease, and 3.18–3.21 for complex MLTCs (**Table 2**).

As in UK Biobank, PAF differences in those with BMI ≥30 did not exceed 1%. In the BMI 25–29.9 group, PAFs were slightly higher, with up to 3.9 percentage point differences between BMI and WC for all-cause mortality, though generally more modest than in UK Biobank (**Table 3**). Similar results were seen in minimally adjusted models (**Supplement eTables 8–9**).

## Discussions

The present analysis, based on two well-characterised prospective cohort studies with granular linkages to multiple diseases, demonstrated that although WC and WHtR have a stronger association with several diseases compared with BMI, the overall gains were modest and for many diseases BMI performed at least as well as the alternative adiposity measures. When considering PAFs, the added value of using WC or WHtR over BMI in explaining MLTCs and mortality risk was surprisingly minimal and statistically non-significant in those with BMI ≥30, <u>supporting the continuing use of BMI alone above this threshold</u> determine overall obesity associated risks. In contrast, some evidence from both cohorts suggests WC or WHtR may explain the risks of MLTCs and mortality than BMI in the overweight range (BMI 25–29.9), challenging the widely held view that these measures are more informative up to BMIs of 35 (as indicated by NICE^11^) or up to 40 as in a recent consensus paper.^4^ In some ways, these new findings make sense, as once BMI is above 30, additional weight gain often leads to parallel gains in central and total adiposity measures.

This work is significant as obesity rates have surged since the 1980s, while longevity has increased - even among those with chronic conditions. As a result, more people are living longer with higher adiposity, contributing to rising MLTCs in high-income countries.^12^ These demographic shifts, along with advances in weight-loss treatments, have intensified research into optimal ways to assess obesity, with growing support for using central adiposity measures.

The present study based on near identical findings in two independent cohorts therefore has clinical implications given that some regulatory authorities – such as the UK NICE – use PAFs to decide which patients should gain preference for weight loss medications.^11^ The present findings suggest that the continued reliance on BMI ≥30 in many clinical guidelines, particularly as a criterion for deciding on interventions targeting weight, likely remains justified. Any shift towards greater use of central adiposity measures should therefore be weighted carefully against their incremental benefits and implementation challenges. Such measures might indeed be useful in people with BMI levels ≥30 but more research is needed to confirm the present findings.

The debate around how best to diagnose obesity has increased in last few years with welcome statements around placing greater emphasis on the signs and symptoms of obesity.^4^ However, debates remain over when BMI alone is sufficient as a starting point to diagnose obesity,^4^ or at which BMI ranges should it be supplemented with additional measures of adiposity such as WC and WHtR.^4,13^ These important questions motivated the present analyses. The findings show that there are some outcomes, such as diabetes, where WC and WHtR are better predictors than BMI. However, even for these outcomes, BMI appeared still a reasonably strong predictor and for many other outcomes, BMI was as good as the alternative measures.

Notably, to gauge a wider clinical context, health care professionals rarely rely on BMI alone to assess many diseases, including incident type 2 diabetes, risks. Instead, they consider several risk factors, or where they exist, risk scores. In the context of obesity, commonly assessed factors include age, ethnicity, lipids, blood pressure, liver function tests, glycaemia markers, and familial histories of disease. These additional factors likely capture most of the extra risk associated with central obesity (potentially by reflecting elements of ectopic fat), limiting the additional predictive value of central over general obesity.^14^ This may partially explain why WC and waist-hip ratio did not provide additional predictive value beyond BMI for cardiovascular events in a large individual participant meta-analysis of 58 cohorts.^15^ Similarly, adding WC to the Framingham Risk Score did not improve prediction of fatal and nonfatal CVD or all-cause mortality.^16^

An additional consideration is that WC is rarely measured in routine clinical practice, and many clinicians are reluctant to incorporate it into patient assessments.^17^ The measurement error for WC is also greater than for weight or height used for the assessment of BMI.^15^ These factors and the new data herein collectively suggest that using BMI as a primary measure of obesity in the general population remains a reasonable and pragmatic approach in those with BMI ≥30. More accurate but also more complex imaging measures, such as DEXA or MRI, are substantially more resource-intensive in terms of costs and time, making them impractical for large-scale use.

This study has several strengths and limitations. Firstly, although the findings were validated in two cohorts using very granular incident outcome data across many outcomes, neither the UK Biobank nor the Whitehall study is representative of the UK population, and therefore the findings, especially absolute risks, are not directly generalisable. However, previous research has shown that exposure-outcome associations can be generalisable in non-representative cohort studies, such as the Whitehall study, despite differences in risk factor frequencies and disease incidence.^18^ UK Biobank data has also been instrumental in many guidelines. Although the findings regarding the relative performance of the alternative obesity measures might be generalisable as well, replications in representative population-based studies are warranted. Secondly, this is an observational data where causality cannot be established, though this work did not seek to look at causality but rather aimed to compare the value of using WC or WHtR versus BMI in predicting MLTCs and mortality. Residual confounding cannot be ruled out. However, there is no evidence to suggest that confounding differed between obesity measures and, therefore, the relative performance should not be affected – notably, the findings were almost identical in both minimally and more comprehensive adjusted models. Thirdly, the PAF statistics serve as a proxy for assessing the potential impact of adopting alternative obesity measures. When comparing measures, PAFs should be interpreted in relative terms, rather than as absolute estimates, which are likely subject to residual confounding. Furthermore, outcomes were ascertained using linked hospital admission and death records, and therefore likely milder cases may have been missed. However, this misclassification is likely to be non-differential and to affect all obesity measures similarly, so the overall conclusions of the study should not be affected. Lastly, the data are mainly from white people; validation of this work needs to be conducted in non-white populations.

In conclusion, given its superior measurement accuracy (lower intra- and inter-observer variability), health care provider preference, and decent predictive performance for a range of obesity-related conditions, MLTCs and mortality compared to WC and WHtR, BMI appears to be the best current simple measure of adiposity for clinical practice in those with BMI≥30. More research is needed to validate these important and surprising findings. Further work is also needed to examine the value of using WC or WHtR over BMI in predicting MLTCs and mortality among individuals in the overweight category.

## Data Availability

Researchers registered with the UK Biobank can apply for access to the individual-level data by completing an application. This must include a summary of the research plan, data fields required, any new data or variables that will be generated and payment to cover the incremental costs of servicing an application (https://www.ukbiobank.ac.uk/enable-your-research/apply-for-access).
Individual-level data from the Whitehall study are also available for sharing within the scientific community. Bona fide researchers interested in accessing the pseudonymized data can apply through Dementias Platform UK (https://www.dementiasplatform.uk) or the Whitehall Scientific Committee (https://www.ucl.ac.uk/epidemiology-health-care/research/epidemiology-and-publichealth/research/whitehall-ii/data-sharing).

## Author Contributions

NS had the original idea refined by FH and MK. FH and JP conducted the analysis. All authors wrote the paper and revised it for intellectual content.

## Data Access, Responsibility, and Analysis

FH and JP had full access to all the data in the study and take responsibility for the integrity of the data and the accuracy of the data analysis.

## Data Sharing Statement

Researchers registered with the UK Biobank can apply for access to the individual-level data by completing an application. This must include a summary of the research plan, data fields required, any new data or variables that will be generated and payment to cover the incremental costs of servicing an application (https://www.ukbiobank.ac.uk/enable-your-research/apply-for-access).

Individual-level data from the Whitehall study are also available for sharing within the scientific community. Bona fide researchers interested in accessing the pseudonymized data can apply through Dementias Platform UK (https://www.dementiasplatform.uk) or the Whitehall Scientific Committee (https://www.ucl.ac.uk/epidemiology-health-care/research/epidemiology-and-publichealth/research/whitehall-ii/data-sharing).

