## Supplement for "Waist, waist-height-ratio vs body mass index and the risks of multiple diseases: a cohort study with replication"

Prof. Mika Kivimäki, FMedSci, Prof. Naveed Sattar, FMedSci

**SUPPLEMENT**

Supplement eTable 1. Cut-points used to define overweight and obesity

| | Female | | | Male | | |
|---|---|---|---|---|---|---|
| | **BMI=25** | **BMI=30** | **BMI=35** | **BMI=25** | **BMI=30** | **BMI=35** |
| Historic | | | | | | |
| WC (Lean et al, 1995) | 80 | 88 | 88 | 94 | 102 | 102 |
| Contemporary (2005-2019) | | | | | | |
| WC | 83 | 96 | 96 | 91 | 106 | 106 |
| WHtR | 0.52 | 0.60 | 0.60 | 0.52 | 0.61 | 0.61 |
| UK Biobank (2007-2010) | | | | | | |
| WC | 80 | 93 | 104 | 89 | 103 | 116 |
| WHtR | 0.49 | 0.57 | 0.64 | 0.51 | 0.59 | 0.66 |
| Whitehall (1991-1994) | | | | | | |
| WC | 75 | 88 | 100 | 88 | 103 | 115 |
| WHtR | 0.47 | 0.55 | 0.62 | 0.50 | 0.58 | 0.66 |

Supplement eTable 2. Associations of obesity with outcomes

| | | BMI | | | | WC (UKB) | | | | WHtR (UKB) | | | |
|---|---|---|---|---|---|---|---|---|---|---|---|---|---|
| | | n | event | HR (95% CI) | P-value | n | event | HR (95% CI) | P-value | n | event | HR (95% CI) | P-value |
| 16 | Diabetes mellitus | 118336 | 21367 | 6.79 (6.56-7.03) | < 0.0001 | 124369 | 23014 | 7.87 (7.58-8.18) | < 0.0001 | 120413 | 23122 | 8.87 (8.53-9.22) | < 0.0001 |
| 29 | Sleep disorders | 120522 | 5807 | 6.25 (5.85-6.67) | < 0.0001 | 126794 | 5939 | 6.38 (5.95-6.84) | < 0.0001 | 122981 | 5757 | 6.47 (6.05-6.92) | < 0.0001 |
| 60 | Gout | 121484 | 4924 | 4.60 (4.31-4.92) | < 0.0001 | 127814 | 5253 | 4.57 (4.25-4.92) | < 0.0001 | 123939 | 5056 | 4.65 (4.33-4.99) | < 0.0001 |
| 0.3 | **Complex multimorbidity (4th disease)** | 119945 | 32402 | 3.19 (3.13-3.26) | < 0.0001 | 126143 | 35230 | 3.45 (3.38-3.53) | < 0.0001 | 122232 | 35389 | 3.60 (3.52-3.68) | < 0.0001 |
| 33 | Hypertension | 109779 | 47696 | 2.79 (2.75-2.84) | < 0.0001 | 115391 | 51327 | 2.86 (2.81-2.90) | < 0.0001 | 111328 | 51052 | 3.03 (2.98-3.08) | < 0.0001 |
| 15 | **Endocrine diseases** | 110339 | 52914 | 2.78 (2.74-2.82) | < 0.0001 | 116068 | 56059 | 2.83 (2.79-2.87) | < 0.0001 | 111939 | 55456 | 2.94 (2.90-2.99) | < 0.0001 |
| 0.2 | **Third disease** | 116534 | 44389 | 2.78 (2.73-2.83) | < 0.0001 | 122486 | 48022 | 2.93 (2.88-2.98) | < 0.0001 | 118472 | 47822 | 3.01 (2.96-3.07) | < 0.0001 |
| 53 | Liver diseases | 121278 | 6547 | 2.53 (2.42-2.64) | < 0.0001 | 127559 | 7099 | 2.86 (2.73-3.00) | < 0.0001 | 123652 | 7072 | 3.04 (2.90-3.19) | < 0.0001 |
| 39 | Heart failure | 121083 | 8054 | 2.45 (2.35-2.55) | < 0.0001 | 127378 | 8829 | 2.57 (2.46-2.69) | < 0.0001 | 123474 | 8882 | 2.57 (2.46-2.68) | < 0.0001 |
| 61 | Osteoarthritis | 114910 | 29546 | 2.40 (2.35-2.44) | < 0.0001 | 121029 | 30639 | 2.22 (2.18-2.27) | < 0.0001 | 117075 | 30438 | 2.20 (2.16-2.25) | < 0.0001 |
| 0.1 | **Simple multimorbidity (2nd disease)** | 108016 | 55569 | 2.38 (2.35-2.41) | < 0.0001 | 113373 | 59830 | 2.44 (2.41-2.48) | < 0.0001 | 109304 | 58913 | 2.49 (2.45-2.52) | < 0.0001 |
| 66 | Renal failure | 121403 | 16099 | 2.35 (2.28-2.41) | < 0.0001 | 127677 | 17679 | 2.46 (2.39-2.53) | < 0.0001 | 123792 | 17890 | 2.53 (2.46-2.60) | < 0.0001 |
| 35 | *Angina pectoris* | 118050 | 8715 | 2.24 (2.16-2.33) | < 0.0001 | 124112 | 9428 | 2.25 (2.17-2.34) | < 0.0001 | 120110 | 9562 | 2.54 (2.44-2.64) | < 0.0001 |
| 55 | Pancreatitis | 121540 | 1219 | 2.23 (2.02-2.46) | < 0.0001 | 127840 | 1328 | 2.62 (2.35-2.91) | < 0.0001 | 123967 | 1310 | 2.69 (2.42-2.99) | < 0.0001 |
| 37 | Pulmonary embolism | 121326 | 3365 | 2.19 (2.06-2.32) | < 0.0001 | 127607 | 3636 | 2.36 (2.21-2.51) | < 0.0001 | 123788 | 3496 | 2.08 (1.96-2.21) | < 0.0001 |
| 32 | **Circulatory diseases** | 101444 | 55033 | 2.07 (2.04-2.09) | < 0.0001 | 106542 | 59417 | 2.11 (2.08-2.14) | < 0.0001 | 102783 | 58454 | 2.15 (2.12-2.18) | < 0.0001 |
| 57 | Eczema and skin infections | 118722 | 9011 | 2.05 (1.98-2.12) | < 0.0001 | 124922 | 9774 | 2.18 (2.10-2.26) | < 0.0001 | 121123 | 9472 | 2.16 (2.09-2.24) | < 0.0001 |
| 34 | Ischaemic heart disease | 116327 | 16759 | 2.00 (1.95-2.05) | < 0.0001 | 122269 | 18304 | 2.03 (1.98-2.08) | < 0.0001 | 118200 | 18277 | 2.18 (2.12-2.24) | < 0.0001 |
| 45 | Deep vein thrombosis | 120942 | 2760 | 1.90 (1.79-2.02) | < 0.0001 | 127178 | 2941 | 1.98 (1.86-2.12) | < 0.0001 | 123377 | 2814 | 1.79 (1.68-1.91) | < 0.0001 |
| 62 | Sciatica | 120505 | 5107 | 1.86 (1.78-1.94) | < 0.0001 | 126797 | 5326 | 1.80 (1.72-1.88) | < 0.0001 | 123011 | 5266 | 1.78 (1.71-1.87) | < 0.0001 |
| 10 | Kidney cancer | 121866 | 765 | 1.85 (1.64-2.08) | < 0.0001 | 128206 | 837 | 1.95 (1.72-2.21) | < 0.0001 | 124334 | 820 | 1.91 (1.69-2.16) | < 0.0001 |
| 63 | Backpain | 119048 | 9101 | 1.81 (1.75-1.87) | < 0.0001 | 125263 | 9680 | 1.82 (1.76-1.88) | < 0.0001 | 121410 | 9665 | 1.86 (1.80-1.92) | < 0.0001 |
| 21 | Mood disorders | 120312 | 10227 | 1.79 (1.74-1.85) | < 0.0001 | 126549 | 10998 | 1.97 (1.91-2.04) | < 0.0001 | 122670 | 10879 | 2.01 (1.95-2.07) | < 0.0001 |
| 49 | Asthma | 117728 | 12513 | 1.79 (1.74-1.84) | < 0.0001 | 123727 | 13458 | 2.00 (1.94-2.06) | < 0.0001 | 119740 | 13468 | 2.05 (1.99-2.11) | < 0.0001 |
| 38 | Arrhythmias | 119804 | 15201 | 1.74 (1.69-1.78) | < 0.0001 | 125892 | 16855 | 1.82 (1.77-1.87) | < 0.0001 | 122132 | 16070 | 1.60 (1.56-1.64) | < 0.0001 |
| 64 | Soft tissue diseases | 115140 | 15702 | 1.73 (1.69-1.77) | < 0.0001 | 121211 | 16352 | 1.70 (1.66-1.74) | < 0.0001 | 117409 | 16157 | 1.72 (1.67-1.76) | < 0.0001 |
| 36 | *Myocardial infarction* | 120092 | 4555 | 1.73 (1.65-1.81) | < 0.0001 | 126284 | 4970 | 1.72 (1.64-1.81) | < 0.0001 | 122342 | 5021 | 1.94 (1.85-2.04) | < 0.0001 |
| 58 | **Musculoskeletal diseases** | 103567 | 45213 | 1.71 (1.69-1.73) | < 0.0001 | 109188 | 47931 | 1.67 (1.65-1.70) | < 0.0001 | 105506 | 47273 | 1.68 (1.65-1.70) | < 0.0001 |
| 23 | **Neurological disorders** | 114226 | 21683 | 1.69 (1.66-1.73) | < 0.0001 | 120293 | 23173 | 1.71 (1.68-1.75) | < 0.0001 | 116360 | 22972 | 1.77 (1.74-1.81) | < 0.0001 |
| 54 | Alcohol-related liver diseases | 121881 | 660 | 1.69 (1.49-1.91) | < 0.0001 | 128215 | 777 | 2.26 (1.96-2.60) | < 0.0001 | 124320 | 787 | 2.79 (2.42-3.23) | < 0.0001 |
| 67 | **Circulatory and respiratory symptoms** | 111022 | 21308 | 1.57 (1.54-1.60) | < 0.0001 | 116786 | 23051 | 1.64 (1.60-1.67) | < 0.0001 | 112907 | 22808 | 1.65 (1.62-1.69) | < 0.0001 |
| 65 | **Genitourinary diseases** | 101913 | 32241 | 1.54 (1.51-1.57) | < 0.0001 | 107517 | 35095 | 1.63 (1.61-1.66) | < 0.0001 | 103469 | 34544 | 1.64 (1.61-1.67) | < 0.0001 |
| 2 | Bacterial infections | 120550 | 12966 | 1.51 (1.47-1.56) | < 0.0001 | 126763 | 14169 | 1.65 (1.60-1.69) | < 0.0001 | 122856 | 14266 | 1.69 (1.65-1.74) | < 0.0001 |
| 47 | Respiratory tract infections | 120869 | 10996 | 1.51 (1.47-1.56) | < 0.0001 | 127102 | 12557 | 1.75 (1.69-1.80) | < 0.0001 | 123212 | 12637 | 1.81 (1.75-1.87) | < 0.0001 |
| 1 | **Infectious diseases** | 117400 | 20166 | 1.49 (1.46-1.53) | < 0.0001 | 123502 | 22005 | 1.60 (1.57-1.64) | < 0.0001 | 119511 | 21971 | 1.64 (1.61-1.68) | < 0.0001 |
| 14 | Anaemia | 119031 | 15010 | 1.49 (1.45-1.52) | < 0.0001 | 125205 | 16245 | 1.58 (1.54-1.62) | < 0.0001 | 121242 | 16550 | 1.67 (1.63-1.71) | < 0.0001 |
| 46 | **Respiratory diseases** | 112313 | 31280 | 1.48 (1.46-1.51) | < 0.0001 | 117977 | 34459 | 1.63 (1.60-1.66) | < 0.0001 | 114115 | 34334 | 1.67 (1.65-1.70) | < 0.0001 |
| 43 | *Cerebral infarction* | 121342 | 3218 | 1.48 (1.40-1.56) | < 0.0001 | 127645 | 3632 | 1.55 (1.46-1.64) | < 0.0001 | 123744 | 3649 | 1.59 (1.50-1.68) | < 0.0001 |
| 44 | Arteriosclerosis | 121812 | 1267 | 1.48 (1.36-1.62) | < 0.0001 | 128139 | 1489 | 1.63 (1.48-1.79) | < 0.0001 | 124236 | 1532 | 1.86 (1.69-2.05) | < 0.0001 |
| 70 | **Poisoning** | 121084 | 1219 | 1.48 (1.36-1.61) | < 0.0001 | 127377 | 1328 | 1.61 (1.48-1.76) | < 0.0001 | 123505 | 1305 | 1.70 (1.56-1.86) | < 0.0001 |
| 13 | **Blood diseases** | 118569 | 16756 | 1.45 (1.42-1.48) | < 0.0001 | 124679 | 18100 | 1.54 (1.50-1.58) | < 0.0001 | 120724 | 18356 | 1.61 (1.57-1.65) | < 0.0001 |
| 56 | **Skin diseases** | 113576 | 17798 | 1.44 (1.40-1.47) | < 0.0001 | 119471 | 19566 | 1.52 (1.48-1.55) | < 0.0001 | 115772 | 18951 | 1.47 (1.44-1.51) | < 0.0001 |
| 17 | **Mental and behavioural disorders** | 118719 | 23311 | 1.43 (1.40-1.45) | < 0.0001 | 124752 | 25711 | 1.54 (1.51-1.57) | < 0.0001 | 120832 | 25555 | 1.60 (1.57-1.63) | < 0.0001 |
| 22 | Neurotic disorders | 121215 | 7903 | 1.38 (1.33-1.42) | < 0.0001 | 127517 | 8603 | 1.52 (1.47-1.57) | < 0.0001 | 123626 | 8580 | 1.53 (1.48-1.59) | < 0.0001 |
| 28 | Transient ischaemic attack | 121553 | 1385 | 1.35 (1.25-1.46) | < 0.0001 | 127874 | 1561 | 1.37 (1.26-1.48) | < 0.0001 | 123966 | 1569 | 1.46 (1.35-1.59) | < 0.0001 |
| 50 | **Digestive system diseases** | 95804 | 47367 | 1.34 (1.32-1.36) | < 0.0001 | 100308 | 50807 | 1.41 (1.39-1.43) | < 0.0001 | 96609 | 49761 | 1.44 (1.42-1.45) | < 0.0001 |
| 20 | Psychotic disorders | 121749 | 516 | 1.34 (1.18-1.52) | 0.003 | 128060 | 619 | 1.65 (1.44-1.88) | < 0.0001 | 124186 | 622 | 1.81 (1.58-2.07) | < 0.0001 |
| 40 | Cerebrovascular diseases | 120769 | 6744 | 1.33 (1.28-1.38) | < 0.0001 | 127039 | 7706 | 1.43 (1.38-1.49) | < 0.0001 | 123121 | 7824 | 1.50 (1.45-1.56) | < 0.0001 |
| 48 | Chronic obstructive pulmonary disease | 121080 | 7799 | 1.33 (1.28-1.37) | < 0.0001 | 127307 | 9265 | 1.57 (1.52-1.63) | < 0.0001 | 123376 | 9504 | 1.70 (1.64-1.76) | < 0.0001 |
| 0 | **All-cause mortality** | 122001 | 13702 | 1.31 (1.28-1.35) | < 0.0001 | 128358 | 15696 | 1.42 (1.38-1.46) | < 0.0001 | 124472 | 15872 | 1.47 (1.43-1.51) | < 0.0001 |
| 59 | Rheumatoid arthritis | 118415 | 18570 | 1.31 (1.28-1.34) | < 0.0001 | 124569 | 20238 | 1.36 (1.33-1.39) | < 0.0001 | 120554 | 20737 | 1.41 (1.38-1.44) | < 0.0001 |
| 41 | Stroke | 121101 | 3955 | 1.31 (1.25-1.37) | < 0.0001 | 127382 | 4490 | 1.38 (1.31-1.45) | < 0.0001 | 123482 | 4518 | 1.41 (1.35-1.48) | < 0.0001 |
| 19 | Substance use disorders | 120940 | 8876 | 1.30 (1.26-1.34) | < 0.0001 | 127123 | 10124 | 1.40 (1.36-1.45) | < 0.0001 | 123211 | 9973 | 1.49 (1.44-1.54) | < 0.0001 |
| 73 | **Self-harm** | 121309 | 625 | 1.30 (1.16-1.46) | 0.002 | 127628 | 686 | 1.41 (1.26-1.59) | < 0.0001 | 123747 | 662 | 1.49 (1.33-1.67) | < 0.0001 |
| 30 | **Eye diseases** | 116484 | 23717 | 1.24 (1.21-1.26) | < 0.0001 | 122301 | 26375 | 1.28 (1.26-1.31) | < 0.0001 | 118330 | 26759 | 1.31 (1.29-1.34) | < 0.0001 |
| 26 | Epilepsy | 121440 | 1655 | 1.24 (1.15-1.33) | < 0.0001 | 127763 | 1856 | 1.37 (1.27-1.47) | < 0.0001 | 123867 | 1857 | 1.47 (1.36-1.58) | < 0.0001 |
| 3 | Viral infections | 121161 | 1863 | 1.22 (1.14-1.30) | < 0.0001 | 127479 | 1985 | 1.23 (1.15-1.31) | < 0.0001 | 123598 | 1989 | 1.26 (1.18-1.35) | < 0.0001 |
| 5 | Colorectal cancer | 121486 | 2309 | 1.21 (1.14-1.28) | < 0.0001 | 127739 | 2626 | 1.29 (1.21-1.37) | < 0.0001 | 123872 | 2545 | 1.24 (1.17-1.32) | < 0.0001 |
| 31 | **Ear diseases** | 121334 | 945 | 1.21 (1.10-1.33) | 0.01 | 127682 | 1024 | 1.24 (1.13-1.36) | 0.004 | 123782 | 1018 | 1.30 (1.18-1.43) | < 0.0001 |
| 68 | **Digestive and abdominal symptoms** | 111863 | 22042 | 1.20 (1.18-1.23) | < 0.0001 | 117703 | 23828 | 1.27 (1.24-1.29) | < 0.0001 | 113697 | 23712 | 1.29 (1.27-1.32) | < 0.0001 |
| 52 | Inflammatory bowel disease | 118489 | 5575 | 1.20 (1.15-1.24) | < 0.0001 | 124584 | 6146 | 1.31 (1.26-1.36) | < 0.0001 | 120646 | 6103 | 1.34 (1.29-1.39) | < 0.0001 |
| 8 | Breast cancer (Female) | 62535 | 3171 | 1.15 (1.10-1.21) | < 0.0001 | 64015 | 3322 | 1.22 (1.16-1.28) | < 0.0001 | 65942 | 3323 | 1.15 (1.09-1.20) | < 0.0001 |
| 27 | Headaches | 121470 | 2004 | 1.15 (1.08-1.23) | 0.002 | 127811 | 2037 | 1.18 (1.11-1.25) | 0.0001 | 123920 | 2006 | 1.15 (1.08-1.22) | 0.006 |
| 72 | **Fall** | 118854 | 10306 | 1.12 (1.09-1.15) | < 0.0001 | 124919 | 11605 | 1.22 (1.18-1.25) | < 0.0001 | 121056 | 11586 | 1.20 (1.17-1.24) | < 0.0001 |
| 4 | **Cancers** | 116775 | 20966 | 1.07 (1.05-1.09) | < 0.0001 | 122554 | 23432 | 1.13 (1.10-1.15) | < 0.0001 | 118703 | 22644 | 1.08 (1.05-1.10) | < 0.0001 |
| 69 | **Injury** | 115233 | 14186 | 1.06 (1.04-1.09) | < 0.0001 | 121175 | 15702 | 1.13 (1.11-1.16) | < 0.0001 | 117503 | 15372 | 1.12 (1.09-1.14) | < 0.0001 |
| 9 | Prostate cancer (Male) | 57328 | 3135 | 0.92 (0.88-0.97) | 0.02 | 61993 | 3728 | 0.95 (0.91-1.01) | 1.00 | 56135 | 3374 | 0.90 (0.85-0.94) | 0.0007 |
| 7 | Melanoma | 120796 | 5370 | 0.85 (0.82-0.88) | < 0.0001 | 126999 | 6116 | 0.89 (0.86-0.92) | < 0.0001 | 123122 | 5806 | 0.83 (0.80-0.86) | < 0.0001 |
| 71 | **Road accidents** | 121046 | 1279 | 0.74 (0.69-0.80) | < 0.0001 | 127382 | 1360 | 0.74 (0.69-0.79) | < 0.0001 | 123535 | 1266 | 0.71 (0.66-0.76) | < 0.0001 |

Supplement eTable 3. Associations of altneriave definitions of obesity with outcomes

| | | BMI | | | | WC (historic) | | | | WC (HES-SHS) | | | | WHtR (HES-SHS) | | | |
|---|---|---|---|---|---|---|---|---|---|---|---|---|---|---|---|---|---|
| | | n | event | HR (95% CI) | P-value | n | event | HR (95% CI) | P-value | n | event | HR (95% CI) | P-value | n | event | HR (95% CI) | P-value |
| 16 | Diabetes mellitus | 118336 | 21367 | 6.79 (6.56-7.03) | < 0.0001 | 163798 | 26052 | 5.60 (5.43-5.77) | < 0.0001 | 92188 | 19215 | 7.92 (7.67-8.19) | < 0.0001 | 79920 | 18262 | 9.00 (8.71-9.30) | < 0.0001 |
| 29 | Sleep disorders | 120522 | 5807 | 6.25 (5.85-6.67) | < 0.0001 | 166542 | 6468 | 4.91 (4.64-5.20) | < 0.0001 | 94299 | 5115 | 6.83 (6.43-7.27) | < 0.0001 | 82011 | 4791 | 7.14 (6.73-7.57) | < 0.0001 |
| 60 | Gout | 121484 | 4924 | 4.60 (4.31-4.92) | < 0.0001 | 167646 | 5779 | 3.23 (3.06-3.41) | < 0.0001 | 95161 | 4238 | 4.48 (4.21-4.78) | < 0.0001 | 82787 | 3896 | 4.73 (4.44-5.04) | < 0.0001 |
| 0.3 | **Complex multimorbidity (4th disease)** | 119945 | 32402 | 3.19 (3.13-3.26) | < 0.0001 | 165793 | 41485 | 2.84 (2.79-2.89) | < 0.0001 | 93710 | 28530 | 3.57 (3.50-3.65) | < 0.0001 | 81352 | 26571 | 3.77 (3.70-3.85) | < 0.0001 |
| 33 | Hypertension | 109779 | 47696 | 2.79 (2.75-2.84) | < 0.0001 | 152872 | 62936 | 2.40 (2.37-2.43) | < 0.0001 | 85148 | 39856 | 2.88 (2.84-2.93) | < 0.0001 | 73296 | 36334 | 3.06 (3.02-3.11) | < 0.0001 |
| 15 | **Endocrine diseases** | 110339 | 52914 | 2.78 (2.74-2.82) | < 0.0001 | 153621 | 68717 | 2.42 (2.39-2.45) | < 0.0001 | 85596 | 43985 | 2.93 (2.89-2.97) | < 0.0001 | 73754 | 39929 | 3.10 (3.05-3.14) | < 0.0001 |
| 0.2 | **Third disease** | 116534 | 44389 | 2.78 (2.73-2.83) | < 0.0001 | 161593 | 58005 | 2.48 (2.44-2.52) | < 0.0001 | 90686 | 38062 | 3.03 (2.98-3.08) | < 0.0001 | 78482 | 34886 | 3.16 (3.11-3.21) | < 0.0001 |
| 53 | Liver diseases | 121278 | 6547 | 2.53 (2.42-2.64) | < 0.0001 | 167272 | 8451 | 2.49 (2.40-2.60) | < 0.0001 | 94961 | 5755 | 2.97 (2.84-3.11) | < 0.0001 | 82592 | 5298 | 3.09 (2.96-3.23) | < 0.0001 |
| 39 | Heart failure | 121083 | 8054 | 2.45 (2.35-2.55) | < 0.0001 | 167137 | 10103 | 2.24 (2.16-2.32) | < 0.0001 | 94778 | 7256 | 2.75 (2.64-2.86) | < 0.0001 | 82416 | 6928 | 2.89 (2.77-3.00) | < 0.0001 |
| 61 | Osteoarthritis | 114910 | 29546 | 2.40 (2.35-2.44) | < 0.0001 | 159263 | 38625 | 1.97 (1.94-2.00) | < 0.0001 | 89689 | 23969 | 2.26 (2.21-2.30) | < 0.0001 | 77783 | 21562 | 2.26 (2.21-2.30) | < 0.0001 |
| 0.1 | **Simple multimorbidity (2nd disease)** | 108016 | 55569 | 2.38 (2.35-2.41) | < 0.0001 | 150710 | 74413 | 2.12 (2.09-2.14) | < 0.0001 | 83394 | 46171 | 2.53 (2.49-2.56) | < 0.0001 | 71672 | 41429 | 2.59 (2.56-2.63) | < 0.0001 |
| 66 | Renal failure | 121403 | 16099 | 2.35 (2.28-2.41) | < 0.0001 | 167466 | 21111 | 2.15 (2.09-2.20) | < 0.0001 | 95066 | 14233 | 2.55 (2.48-2.62) | < 0.0001 | 82683 | 13432 | 2.68 (2.61-2.75) | < 0.0001 |
| 35 | *Angina pectoris* | 118050 | 8715 | 2.24 (2.16-2.33) | < 0.0001 | 163298 | 11227 | 1.92 (1.86-1.99) | < 0.0001 | 92280 | 7356 | 2.22 (2.14-2.31) | < 0.0001 | 79966 | 6994 | 2.48 (2.39-2.57) | < 0.0001 |
| 55 | Pancreatitis | 121540 | 1219 | 2.23 (2.02-2.46) | < 0.0001 | 167623 | 1604 | 2.31 (2.11-2.53) | < 0.0001 | 95206 | 1074 | 2.61 (2.37-2.88) | < 0.0001 | 82839 | 949 | 2.59 (2.34-2.86) | < 0.0001 |
| 37 | Pulmonary embolism | 121326 | 3365 | 2.19 (2.06-2.32) | < 0.0001 | 167361 | 4453 | 2.08 (1.97-2.19) | < 0.0001 | 95003 | 2880 | 2.33 (2.20-2.47) | < 0.0001 | 82692 | 2532 | 2.10 (1.98-2.22) | < 0.0001 |
| 32 | **Circulatory diseases** | 101444 | 55033 | 2.07 (2.04-2.09) | < 0.0001 | 141400 | 74344 | 1.87 (1.85-1.90) | < 0.0001 | 78591 | 45482 | 2.17 (2.14-2.20) | < 0.0001 | 67613 | 40693 | 2.23 (2.20-2.26) | < 0.0001 |
| 57 | Eczema and skin infections | 118722 | 9011 | 2.05 (1.98-2.12) | < 0.0001 | 164217 | 11576 | 1.98 (1.92-2.05) | < 0.0001 | 92770 | 7916 | 2.35 (2.27-2.44) | < 0.0001 | 80656 | 7186 | 2.38 (2.30-2.46) | < 0.0001 |
| 34 | Ischaemic heart disease | 116327 | 16759 | 2.00 (1.95-2.05) | < 0.0001 | 161135 | 21790 | 1.79 (1.75-1.83) | < 0.0001 | 90838 | 14211 | 2.05 (2.00-2.10) | < 0.0001 | 78576 | 13246 | 2.22 (2.16-2.28) | < 0.0001 |
| 45 | Deep vein thrombosis | 120942 | 2760 | 1.90 (1.79-2.02) | < 0.0001 | 166887 | 3586 | 1.77 (1.67-1.87) | < 0.0001 | 94641 | 2334 | 2.05 (1.93-2.18) | < 0.0001 | 82378 | 2027 | 1.82 (1.71-1.94) | < 0.0001 |
| 62 | Sciatica | 120505 | 5107 | 1.86 (1.78-1.94) | < 0.0001 | 166260 | 6690 | 1.64 (1.58-1.71) | < 0.0001 | 94403 | 4156 | 1.80 (1.73-1.89) | < 0.0001 | 82169 | 3658 | 1.77 (1.69-1.85) | < 0.0001 |
| 10 | Kidney cancer | 121866 | 765 | 1.85 (1.64-2.08) | < 0.0001 | 168052 | 1008 | 1.72 (1.55-1.90) | < 0.0001 | 95485 | 689 | 1.98 (1.76-2.23) | < 0.0001 | 83106 | 590 | 1.89 (1.67-2.13) | < 0.0001 |
| 63 | Backpain | 119048 | 9101 | 1.81 (1.75-1.87) | < 0.0001 | 164385 | 12147 | 1.69 (1.64-1.74) | < 0.0001 | 93158 | 7607 | 1.87 (1.81-1.93) | < 0.0001 | 81024 | 6913 | 1.90 (1.84-1.97) | < 0.0001 |
| 21 | Mood disorders | 120312 | 10227 | 1.79 (1.74-1.85) | < 0.0001 | 166025 | 13910 | 1.83 (1.78-1.88) | < 0.0001 | 94143 | 8801 | 2.04 (1.98-2.10) | < 0.0001 | 81873 | 7869 | 2.06 (1.99-2.12) | < 0.0001 |
| 49 | Asthma | 117728 | 12513 | 1.79 (1.74-1.84) | < 0.0001 | 162497 | 16913 | 1.77 (1.73-1.82) | < 0.0001 | 91868 | 10601 | 2.02 (1.97-2.08) | < 0.0001 | 79717 | 9659 | 2.12 (2.06-2.18) | < 0.0001 |
| 38 | Arrhythmias | 119804 | 15201 | 1.74 (1.69-1.78) | < 0.0001 | 165317 | 20102 | 1.70 (1.66-1.74) | < 0.0001 | 93665 | 13352 | 1.95 (1.90-2.00) | < 0.0001 | 81517 | 11725 | 1.75 (1.71-1.80) | < 0.0001 |
| 64 | Soft tissue diseases | 115140 | 15702 | 1.73 (1.69-1.77) | < 0.0001 | 159293 | 20608 | 1.55 (1.52-1.58) | < 0.0001 | 90011 | 12824 | 1.75 (1.71-1.80) | < 0.0001 | 78148 | 11478 | 1.77 (1.73-1.81) | < 0.0001 |
| 36 | *Myocardial infarction* | 120092 | 4555 | 1.73 (1.65-1.81) | < 0.0001 | 165880 | 5916 | 1.55 (1.49-1.62) | < 0.0001 | 94005 | 3800 | 1.72 (1.64-1.80) | < 0.0001 | 81643 | 3558 | 1.90 (1.81-1.99) | < 0.0001 |
| 58 | **Musculoskeletal diseases** | 103567 | 45213 | 1.71 (1.69-1.73) | < 0.0001 | 144004 | 61376 | 1.55 (1.53-1.57) | < 0.0001 | 80705 | 36857 | 1.73 (1.70-1.75) | < 0.0001 | 69827 | 32751 | 1.74 (1.72-1.77) | < 0.0001 |
| 23 | **Neurological disorders** | 114226 | 21683 | 1.69 (1.66-1.73) | < 0.0001 | 158266 | 28624 | 1.59 (1.56-1.62) | < 0.0001 | 89148 | 18252 | 1.81 (1.77-1.85) | < 0.0001 | 77210 | 16543 | 1.88 (1.84-1.92) | < 0.0001 |
| 54 | Alcohol-related liver diseases | 121881 | 660 | 1.69 (1.49-1.91) | < 0.0001 | 168065 | 874 | 2.18 (1.94-2.46) | < 0.0001 | 95498 | 628 | 2.52 (2.20-2.89) | < 0.0001 | 83094 | 594 | 2.94 (2.57-3.38) | < 0.0001 |
| 67 | **Circulatory and respiratory symptoms** | 111022 | 21308 | 1.57 (1.54-1.60) | < 0.0001 | 154074 | 28864 | 1.52 (1.50-1.55) | < 0.0001 | 86510 | 17738 | 1.66 (1.63-1.70) | < 0.0001 | 74884 | 16067 | 1.71 (1.67-1.74) | < 0.0001 |
| 65 | **Genitourinary diseases** | 101913 | 32241 | 1.54 (1.51-1.57) | < 0.0001 | 139792 | 43731 | 1.51 (1.49-1.53) | < 0.0001 | 79467 | 27064 | 1.67 (1.64-1.70) | < 0.0001 | 68784 | 24331 | 1.71 (1.68-1.74) | < 0.0001 |
| 2 | Bacterial infections | 120550 | 12966 | 1.51 (1.47-1.56) | < 0.0001 | 166259 | 17575 | 1.54 (1.50-1.58) | < 0.0001 | 94345 | 11301 | 1.72 (1.67-1.76) | < 0.0001 | 82041 | 10357 | 1.78 (1.74-1.83) | < 0.0001 |
| 47 | Respiratory tract infections | 120869 | 10996 | 1.51 (1.47-1.56) | < 0.0001 | 166723 | 14971 | 1.62 (1.58-1.66) | < 0.0001 | 94584 | 10060 | 1.84 (1.78-1.90) | < 0.0001 | 82279 | 9276 | 1.92 (1.87-1.98) | < 0.0001 |
| 1 | **Infectious diseases** | 117400 | 20166 | 1.49 (1.46-1.53) | < 0.0001 | 162139 | 27339 | 1.50 (1.48-1.53) | < 0.0001 | 91744 | 17344 | 1.67 (1.63-1.70) | < 0.0001 | 79620 | 15850 | 1.73 (1.69-1.77) | < 0.0001 |
| 14 | Anaemia | 119031 | 15010 | 1.49 (1.45-1.52) | < 0.0001 | 164183 | 20154 | 1.47 (1.44-1.50) | < 0.0001 | 93127 | 12892 | 1.67 (1.63-1.71) | < 0.0001 | 80877 | 12026 | 1.77 (1.73-1.82) | < 0.0001 |
| 46 | **Respiratory diseases** | 112313 | 31280 | 1.48 (1.46-1.51) | < 0.0001 | 155312 | 42946 | 1.51 (1.49-1.53) | < 0.0001 | 87478 | 26706 | 1.67 (1.65-1.70) | < 0.0001 | 75753 | 24248 | 1.74 (1.71-1.77) | < 0.0001 |
| 43 | *Cerebral infarction* | 121342 | 3218 | 1.48 (1.40-1.56) | < 0.0001 | 167413 | 4428 | 1.45 (1.38-1.52) | < 0.0001 | 95048 | 2796 | 1.58 (1.50-1.67) | < 0.0001 | 82658 | 2632 | 1.68 (1.59-1.77) | < 0.0001 |
| 44 | Arteriosclerosis | 121812 | 1267 | 1.48 (1.36-1.62) | < 0.0001 | 167985 | 1742 | 1.60 (1.47-1.73) | < 0.0001 | 95433 | 1178 | 1.74 (1.59-1.90) | < 0.0001 | 83040 | 1155 | 1.97 (1.80-2.16) | < 0.0001 |
| 70 | **Poisoning** | 121084 | 1219 | 1.48 (1.36-1.61) | < 0.0001 | 167004 | 1649 | 1.54 (1.43-1.67) | < 0.0001 | 94822 | 1033 | 1.66 (1.52-1.81) | < 0.0001 | 82495 | 947 | 1.85 (1.70-2.03) | < 0.0001 |
| 13 | **Blood diseases** | 118569 | 16756 | 1.45 (1.42-1.48) | < 0.0001 | 163489 | 22563 | 1.44 (1.41-1.47) | < 0.0001 | 92728 | 14318 | 1.62 (1.58-1.66) | < 0.0001 | 80522 | 13227 | 1.70 (1.66-1.74) | < 0.0001 |
| 56 | **Skin diseases** | 113576 | 17798 | 1.44 (1.40-1.47) | < 0.0001 | 157162 | 24165 | 1.42 (1.40-1.45) | < 0.0001 | 88608 | 15315 | 1.61 (1.57-1.64) | < 0.0001 | 76947 | 13555 | 1.58 (1.54-1.61) | < 0.0001 |
| 17 | **Mental and behavioural disorders** | 118719 | 23311 | 1.43 (1.40-1.45) | < 0.0001 | 163912 | 32388 | 1.46 (1.44-1.49) | < 0.0001 | 92702 | 20016 | 1.60 (1.57-1.63) | < 0.0001 | 80535 | 18173 | 1.66 (1.63-1.69) | < 0.0001 |
| 22 | Neurotic disorders | 121215 | 7903 | 1.38 (1.33-1.42) | < 0.0001 | 167184 | 11212 | 1.44 (1.40-1.49) | < 0.0001 | 94920 | 6714 | 1.54 (1.49-1.60) | < 0.0001 | 82582 | 6031 | 1.57 (1.52-1.63) | < 0.0001 |
| 28 | Transient ischaemic attack | 121553 | 1385 | 1.35 (1.25-1.46) | < 0.0001 | 167639 | 1951 | 1.31 (1.22-1.40) | < 0.0001 | 95222 | 1165 | 1.39 (1.29-1.51) | < 0.0001 | 82861 | 1082 | 1.45 (1.33-1.57) | < 0.0001 |
| 50 | **Digestive system diseases** | 95804 | 47367 | 1.34 (1.32-1.36) | < 0.0001 | 132625 | 65568 | 1.33 (1.32-1.35) | < 0.0001 | 74273 | 38434 | 1.42 (1.40-1.44) | < 0.0001 | 63908 | 33948 | 1.45 (1.43-1.47) | < 0.0001 |
| 20 | Psychotic disorders | 121749 | 516 | 1.34 (1.18-1.52) | 0.003 | 167893 | 746 | 1.64 (1.45-1.84) | < 0.0001 | 95355 | 500 | 1.81 (1.59-2.07) | < 0.0001 | 82984 | 451 | 1.88 (1.64-2.15) | < 0.0001 |
| 40 | Cerebrovascular diseases | 120769 | 6744 | 1.33 (1.28-1.38) | < 0.0001 | 166651 | 9494 | 1.37 (1.32-1.41) | < 0.0001 | 94582 | 5890 | 1.46 (1.41-1.51) | < 0.0001 | 82209 | 5560 | 1.55 (1.49-1.61) | < 0.0001 |

| | | | | | | | | | | | | | | | | | |
|---|---|---|---|---|---|---|---|---|---|---|---|---|---|---|---|---|---|
| 48 | Chronic obstructive pulmonary disease | 121080 | 7799 | 1.33 (1.28-1.37) | < 0.0001 | 167024 | 11130 | 1.53 (1.48-1.58) | < 0.0001 | 94724 | 7428 | 1.71 (1.65-1.77) | < 0.0001 | 82332 | 7013 | 1.83 (1.77-1.90) | < 0.0001 |
| 0 | **All-cause mortality** | 122001 | 13702 | 1.31 (1.28-1.35) | < 0.0001 | 168235 | 18817 | 1.37 (1.34-1.41) | < 0.0001 | 95604 | 12408 | 1.52 (1.48-1.56) | < 0.0001 | 83209 | 11690 | 1.61 (1.57-1.65) | < 0.0001 |
| 59 | Rheumatoid arthritis | 118415 | 18570 | 1.31 (1.28-1.34) | < 0.0001 | 163429 | 26491 | 1.31 (1.29-1.34) | < 0.0001 | 92603 | 15811 | 1.43 (1.40-1.46) | < 0.0001 | 80365 | 14486 | 1.49 (1.46-1.52) | < 0.0001 |
| 41 | Stroke | 121101 | 3955 | 1.31 (1.25-1.37) | < 0.0001 | 167066 | 5539 | 1.32 (1.27-1.38) | < 0.0001 | 94851 | 3423 | 1.40 (1.33-1.47) | < 0.0001 | 82485 | 3203 | 1.47 (1.40-1.55) | < 0.0001 |
| 19 | Substance use disorders | 120940 | 8876 | 1.30 (1.26-1.34) | < 0.0001 | 166813 | 12483 | 1.36 (1.33-1.40) | < 0.0001 | 94617 | 7816 | 1.45 (1.41-1.50) | < 0.0001 | 82284 | 7049 | 1.52 (1.48-1.57) | < 0.0001 |
| 73 | **Self-harm** | 121309 | 625 | 1.30 (1.16-1.46) | 0.002 | 167316 | 851 | 1.40 (1.27-1.55) | < 0.0001 | 95018 | 535 | 1.48 (1.32-1.66) | < 0.0001 | 82678 | 482 | 1.67 (1.48-1.88) | < 0.0001 |
| 30 | **Eye diseases** | 116484 | 23717 | 1.24 (1.21-1.26) | < 0.0001 | 160495 | 34048 | 1.24 (1.22-1.26) | < 0.0001 | 90935 | 20100 | 1.32 (1.29-1.34) | < 0.0001 | 78881 | 18377 | 1.34 (1.32-1.37) | < 0.0001 |
| 26 | Epilepsy | 121440 | 1655 | 1.24 (1.15-1.33) | < 0.0001 | 167500 | 2294 | 1.32 (1.24-1.41) | < 0.0001 | 95140 | 1431 | 1.42 (1.32-1.52) | < 0.0001 | 82770 | 1340 | 1.53 (1.43-1.65) | < 0.0001 |
| 3 | Viral infections | 121161 | 1863 | 1.22 (1.14-1.30) | < 0.0001 | 167110 | 2518 | 1.18 (1.12-1.26) | < 0.0001 | 94921 | 1529 | 1.24 (1.16-1.33) | < 0.0001 | 82594 | 1366 | 1.28 (1.20-1.37) | < 0.0001 |
| 5 | Colorectal cancer | 121486 | 2309 | 1.21 (1.14-1.28) | < 0.0001 | 167471 | 3290 | 1.26 (1.20-1.33) | < 0.0001 | 95138 | 2010 | 1.34 (1.26-1.43) | < 0.0001 | 82794 | 1767 | 1.29 (1.21-1.37) | < 0.0001 |
| 31 | **Ear diseases** | 121334 | 945 | 1.21 (1.10-1.33) | 0.01 | 167350 | 1309 | 1.24 (1.15-1.35) | < 0.0001 | 95076 | 778 | 1.28 (1.17-1.41) | < 0.0001 | 82734 | 701 | 1.33 (1.21-1.47) | < 0.0001 |
| 68 | **Digestive and abdominal symptoms** | 111863 | 22042 | 1.20 (1.18-1.23) | < 0.0001 | 154268 | 30880 | 1.23 (1.21-1.25) | < 0.0001 | 87383 | 18159 | 1.29 (1.27-1.32) | < 0.0001 | 75774 | 16290 | 1.33 (1.30-1.36) | < 0.0001 |
| 52 | Inflammatory bowel disease | 118489 | 5575 | 1.20 (1.15-1.24) | < 0.0001 | 163365 | 7832 | 1.24 (1.20-1.29) | < 0.0001 | 92656 | 4722 | 1.32 (1.27-1.38) | < 0.0001 | 80495 | 4286 | 1.38 (1.33-1.44) | < 0.0001 |
| 8 | Breast cancer (Female) | 62535 | 3171 | 1.15 (1.10-1.21) | < 0.0001 | 96269 | 4900 | 1.19 (1.14-1.24) | < 0.0001 | 49104 | 2543 | 1.19 (1.13-1.25) | < 0.0001 | 43217 | 2147 | 1.11 (1.05-1.17) | 0.001 |
| 27 | Headaches | 121470 | 2004 | 1.15 (1.08-1.23) | 0.002 | 167511 | 2762 | 1.14 (1.08-1.21) | 0.0005 | 95169 | 1571 | 1.19 (1.12-1.27) | < 0.0001 | 82825 | 1353 | 1.14 (1.06-1.21) | 0.050 |
| 72 | **Fall** | 118854 | 10306 | 1.12 (1.09-1.15) | < 0.0001 | 163839 | 15016 | 1.21 (1.18-1.24) | < 0.0001 | 92891 | 9045 | 1.28 (1.25-1.32) | < 0.0001 | 80794 | 8255 | 1.30 (1.26-1.34) | < 0.0001 |
| 51 | Appendicitis | 121439 | 698 | 1.12 (1.01-1.25) | 1.00 | 167455 | 975 | 1.14 (1.04-1.24) | 1.00 | 95168 | 540 | 1.12 (1.01-1.25) | 1.00 | 82848 | 461 | 1.08 (0.97-1.21) | 1.00 |
| 12 | Leukaemia / Lymphoma | 121601 | 3319 | 1.09 (1.04-1.14) | 0.22 | 167671 | 4729 | 1.14 (1.09-1.19) | < 0.0001 | 95248 | 2863 | 1.20 (1.14-1.26) | < 0.0001 | 82901 | 2480 | 1.12 (1.07-1.18) | 0.004 |
| 4 | **Cancers** | 116775 | 20966 | 1.07 (1.05-1.09) | < 0.0001 | 160526 | 29719 | 1.10 (1.08-1.12) | < 0.0001 | 91194 | 17641 | 1.15 (1.13-1.17) | < 0.0001 | 79309 | 15396 | 1.10 (1.08-1.13) | < 0.0001 |
| 69 | **Injury** | 115233 | 14186 | 1.06 (1.04-1.09) | < 0.0001 | 159238 | 20262 | 1.13 (1.11-1.15) | < 0.0001 | 90088 | 12055 | 1.18 (1.15-1.21) | < 0.0001 | 78396 | 10817 | 1.19 (1.16-1.21) | < 0.0001 |
| 18 | Dementia | 121976 | 2534 | 1.06 (1.00-1.12) | 1.00 | 168199 | 3715 | 1.09 (1.04-1.14) | 0.24 | 95579 | 2257 | 1.19 (1.12-1.26) | < 0.0001 | 83186 | 2257 | 1.31 (1.24-1.39) | < 0.0001 |
| 24 | Parkinson disease | 121946 | 1043 | 1.05 (0.96-1.14) | 1.00 | 168154 | 1556 | 1.16 (1.08-1.26) | 0.03 | 95561 | 948 | 1.21 (1.11-1.32) | 0.009 | 83162 | 849 | 1.19 (1.09-1.30) | 0.06 |
| 6 | Lung cancer | 121927 | 1690 | 0.99 (0.93-1.06) | 1.00 | 168132 | 2593 | 1.12 (1.05-1.19) | 0.07 | 95531 | 1564 | 1.14 (1.07-1.22) | 0.04 | 83143 | 1456 | 1.19 (1.11-1.27) | 0.0005 |
| 11 | Brain cancer | 121982 | 315 | 0.96 (0.82-1.11) | 1.00 | 168208 | 436 | 0.98 (0.85-1.11) | 1.00 | 95588 | 244 | 0.89 (0.76-1.05) | 1.00 | 83193 | 206 | 0.79 (0.67-0.94) | 1.00 |
| 42 | *Intracerebral haemorrhage* | 121747 | 980 | 0.94 (0.86-1.02) | 1.00 | 167868 | 1456 | 1.04 (0.96-1.12) | 1.00 | 95394 | 849 | 1.03 (0.94-1.12) | 1.00 | 83023 | 775 | 1.05 (0.96-1.15) | 1.00 |
| 9 | Prostate cancer (Male) | 57328 | 3135 | 0.92 (0.88-0.97) | 0.02 | 68643 | 4145 | 0.96 (0.92-1.00) | 0.64 | 44735 | 2601 | 0.93 (0.88-0.98) | 0.07 | 38420 | 2285 | 0.90 (0.85-0.95) | 0.001 |
| 7 | Melanoma | 120796 | 5370 | 0.85 (0.82-0.88) | < 0.0001 | 166415 | 7907 | 0.90 (0.87-0.93) | < 0.0001 | 94599 | 4493 | 0.90 (0.87-0.94) | < 0.0001 | 82329 | 3792 | 0.84 (0.81-0.87) | < 0.0001 |
| 25 | Multiple sclerosis | 121830 | 307 | 0.84 (0.73-0.97) | 1.00 | 167960 | 484 | 1.00 (0.88-1.14) | 1.00 | 95447 | 276 | 1.05 (0.90-1.22) | 1.00 | 83067 | 223 | 1.00 (0.85-1.17) | 1.00 |
| 71 | **Road accidents** | 121046 | 1279 | 0.74 (0.69-0.80) | < 0.0001 | 167020 | 1723 | 0.79 (0.75-0.84) | < 0.0001 | 94880 | 994 | 0.77 (0.72-0.84) | < 0.0001 | 82558 | 881 | 0.78 (0.72-0.85) | < 0.0001 |

Supplement eTable 4. Associations of BMI, WC, and WHtR with outcomes

| | | BMI | | | | WC | | | | WHtR | | | |
|---|---|---|---|---|---|---|---|---|---|---|---|---|---|
| | | n | event | HR (95% CI) | P-value | n | event | HR (95% CI) | P-value | n | event | HR (95% CI) | P-value |
| 29 | Sleep disorders | 493230 | 9956 | 1.93 (1.90-1.95) | < 0.0001 | 493230 | 9956 | 2.08 (2.05-2.11) | < 0.0001 | 493230 | 9956 | 2.09 (2.06-2.12) | < 0.0001 |
| 16 | Diabetes mellitus | 489001 | 39361 | 1.81 (1.80-1.82) | < 0.0001 | 489001 | 39361 | 2.01 (1.99-2.02) | < 0.0001 | 489001 | 39361 | 2.05 (2.03-2.06) | < 0.0001 |
| 60 | Gout | 494891 | 10362 | 1.62 (1.60-1.64) | < 0.0001 | 494891 | 10362 | 1.67 (1.64-1.70) | < 0.0001 | 494891 | 10362 | 1.68 (1.65-1.71) | < 0.0001 |
| 0.3 | **Complex multimorbidity (4th disease)** | 492109 | 76063 | 1.59 (1.58-1.60) | < 0.0001 | 492109 | 76063 | 1.66 (1.65-1.67) | < 0.0001 | 492109 | 76063 | 1.68 (1.67-1.69) | < 0.0001 |
| 0.2 | **Third disease** | 484794 | 115949 | 1.52 (1.51-1.53) | < 0.0001 | 484794 | 115949 | 1.57 (1.56-1.58) | < 0.0001 | 484794 | 115949 | 1.58 (1.57-1.59) | < 0.0001 |
| 15 | **Endocrine diseases** | 467853 | 142086 | 1.52 (1.51-1.53) | < 0.0001 | 467853 | 142086 | 1.55 (1.55-1.56) | < 0.0001 | 467853 | 142086 | 1.57 (1.57-1.58) | < 0.0001 |
| 39 | Heart failure | 493856 | 19152 | 1.49 (1.47-1.51) | < 0.0001 | 493856 | 19152 | 1.55 (1.53-1.57) | < 0.0001 | 493856 | 19152 | 1.56 (1.54-1.58) | < 0.0001 |
| 33 | Hypertension | 467784 | 132767 | 1.48 (1.48-1.49) | < 0.0001 | 467784 | 132767 | 1.51 (1.51-1.52) | < 0.0001 | 467784 | 132767 | 1.55 (1.54-1.55) | < 0.0001 |
| 0.1 | **Simple multimorbidity (2nd disease)** | 463681 | 166774 | 1.43 (1.42-1.44) | < 0.0001 | 463681 | 166774 | 1.46 (1.45-1.46) | < 0.0001 | 463681 | 166774 | 1.47 (1.46-1.48) | < 0.0001 |
| 66 | Renal failure | 494325 | 41055 | 1.42 (1.41-1.43) | < 0.0001 | 494325 | 41055 | 1.47 (1.45-1.48) | < 0.0001 | 494325 | 41055 | 1.48 (1.47-1.49) | < 0.0001 |
| 53 | Liver diseases | 494013 | 15857 | 1.42 (1.40-1.44) | < 0.0001 | 494013 | 15857 | 1.51 (1.49-1.53) | < 0.0001 | 494013 | 15857 | 1.53 (1.51-1.55) | < 0.0001 |
| 61 | Osteoarthritis | 478245 | 82728 | 1.41 (1.40-1.42) | < 0.0001 | 478245 | 82728 | 1.39 (1.38-1.40) | < 0.0001 | 478245 | 82728 | 1.39 (1.38-1.40) | < 0.0001 |
| 57 | Eczema and skin infections | 487834 | 23693 | 1.41 (1.39-1.42) | < 0.0001 | 487834 | 23693 | 1.45 (1.44-1.47) | < 0.0001 | 487834 | 23693 | 1.44 (1.43-1.46) | < 0.0001 |
| 37 | Pulmonary embolism | 494283 | 8967 | 1.37 (1.34-1.39) | < 0.0001 | 494283 | 8967 | 1.41 (1.38-1.44) | < 0.0001 | 494283 | 8967 | 1.36 (1.33-1.39) | < 0.0001 |
| 55 | Pancreatitis | 494758 | 3112 | 1.37 (1.33-1.41) | < 0.0001 | 494758 | 3112 | 1.43 (1.38-1.47) | < 0.0001 | 494758 | 3112 | 1.44 (1.39-1.48) | < 0.0001 |
| 32 | **Circulatory diseases** | 435012 | 175304 | 1.35 (1.34-1.36) | < 0.0001 | 435012 | 175304 | 1.37 (1.36-1.37) | < 0.0001 | 435012 | 175304 | 1.38 (1.37-1.38) | < 0.0001 |
| 35 | *Angina pectoris* | 486123 | 23932 | 1.33 (1.32-1.35) | < 0.0001 | 486123 | 23932 | 1.35 (1.33-1.36) | < 0.0001 | 486123 | 23932 | 1.40 (1.38-1.42) | < 0.0001 |
| 34 | Ischaemic heart disease | 481099 | 48187 | 1.31 (1.30-1.32) | < 0.0001 | 481099 | 48187 | 1.33 (1.32-1.34) | < 0.0001 | 481099 | 48187 | 1.36 (1.35-1.37) | < 0.0001 |
| 38 | Arrhythmias | 489334 | 43957 | 1.30 (1.29-1.31) | < 0.0001 | 489334 | 43957 | 1.34 (1.33-1.35) | < 0.0001 | 489334 | 43957 | 1.28 (1.26-1.29) | < 0.0001 |
| 45 | Deep vein thrombosis | 493390 | 7844 | 1.29 (1.26-1.31) | < 0.0001 | 493390 | 7844 | 1.32 (1.30-1.35) | < 0.0001 | 493390 | 7844 | 1.28 (1.25-1.31) | < 0.0001 |
| 23 | **Neurological disorders** | 474833 | 66201 | 1.28 (1.27-1.29) | < 0.0001 | 474833 | 66201 | 1.29 (1.28-1.30) | < 0.0001 | 474833 | 66201 | 1.30 (1.29-1.31) | < 0.0001 |
| 10 | Kidney cancer | 495495 | 2236 | 1.28 (1.23-1.33) | < 0.0001 | 495495 | 2236 | 1.31 (1.26-1.37) | < 0.0001 | 495495 | 2236 | 1.29 (1.24-1.34) | < 0.0001 |
| 21 | Mood disorders | 491472 | 29698 | 1.27 (1.26-1.29) | < 0.0001 | 491472 | 29698 | 1.33 (1.31-1.34) | < 0.0001 | 491472 | 29698 | 1.34 (1.32-1.35) | < 0.0001 |
| 49 | Asthma | 483992 | 37341 | 1.27 (1.26-1.28) | < 0.0001 | 483992 | 37341 | 1.32 (1.31-1.33) | < 0.0001 | 483992 | 37341 | 1.35 (1.34-1.36) | < 0.0001 |
| 63 | Backpain | 487592 | 26519 | 1.26 (1.25-1.27) | < 0.0001 | 487592 | 26519 | 1.29 (1.27-1.30) | < 0.0001 | 487592 | 26519 | 1.29 (1.28-1.31) | < 0.0001 |
| 58 | **Musculoskeletal diseases** | 438286 | 150109 | 1.25 (1.25-1.26) | < 0.0001 | 438286 | 150109 | 1.25 (1.24-1.26) | < 0.0001 | 438286 | 150109 | 1.25 (1.25-1.26) | < 0.0001 |
| 62 | Sciatica | 491506 | 15123 | 1.25 (1.24-1.27) | < 0.0001 | 491506 | 15123 | 1.25 (1.23-1.27) | < 0.0001 | 491506 | 15123 | 1.25 (1.23-1.27) | < 0.0001 |
| 54 | Alcohol-related liver diseases | 495531 | 1701 | 1.25 (1.20-1.30) | < 0.0001 | 495531 | 1701 | 1.42 (1.37-1.49) | < 0.0001 | 495531 | 1701 | 1.50 (1.44-1.56) | < 0.0001 |
| 64 | Soft tissue diseases | 475676 | 48579 | 1.24 (1.23-1.25) | < 0.0001 | 475676 | 48579 | 1.24 (1.23-1.25) | < 0.0001 | 475676 | 48579 | 1.25 (1.24-1.26) | < 0.0001 |
| 36 | *Myocardial infarction* | 490725 | 13883 | 1.23 (1.21-1.25) | < 0.0001 | 490725 | 13883 | 1.24 (1.22-1.26) | < 0.0001 | 490725 | 13883 | 1.28 (1.26-1.30) | < 0.0001 |
| 47 | Respiratory tract infections | 492365 | 32236 | 1.23 (1.21-1.24) | < 0.0001 | 492365 | 32236 | 1.30 (1.28-1.31) | < 0.0001 | 492365 | 32236 | 1.31 (1.30-1.32) | < 0.0001 |
| 2 | Bacterial infections | 491205 | 39490 | 1.21 (1.20-1.22) | < 0.0001 | 491205 | 39490 | 1.26 (1.25-1.27) | < 0.0001 | 491205 | 39490 | 1.27 (1.26-1.28) | < 0.0001 |
| 67 | **Circulatory and respiratory symptoms** | 465131 | 68898 | 1.21 (1.20-1.21) | < 0.0001 | 465131 | 68898 | 1.23 (1.22-1.24) | < 0.0001 | 465131 | 68898 | 1.24 (1.23-1.25) | < 0.0001 |
| 1 | **Infectious diseases** | 481279 | 62795 | 1.20 (1.20-1.21) | < 0.0001 | 481279 | 62795 | 1.24 (1.23-1.25) | < 0.0001 | 481279 | 62795 | 1.25 (1.24-1.26) | < 0.0001 |
| 56 | **Skin diseases** | 469380 | 58065 | 1.20 (1.20-1.21) | < 0.0001 | 469380 | 58065 | 1.23 (1.22-1.24) | < 0.0001 | 469380 | 58065 | 1.22 (1.21-1.23) | < 0.0001 |
| 65 | **Genitourinary diseases** | 424122 | 105547 | 1.20 (1.20-1.21) | < 0.0001 | 424122 | 105547 | 1.23 (1.22-1.24) | < 0.0001 | 424122 | 105547 | 1.23 (1.23-1.24) | < 0.0001 |
| 14 | Anaemia | 485916 | 45783 | 1.20 (1.19-1.21) | < 0.0001 | 485916 | 45783 | 1.23 (1.22-1.24) | < 0.0001 | 485916 | 45783 | 1.25 (1.24-1.26) | < 0.0001 |
| 44 | Arteriosclerosis | 495381 | 3744 | 1.20 (1.16-1.24) | < 0.0001 | 495381 | 3744 | 1.28 (1.24-1.32) | < 0.0001 | 495381 | 3744 | 1.32 (1.28-1.36) | < 0.0001 |
| 46 | **Respiratory diseases** | 465724 | 101286 | 1.19 (1.18-1.20) | < 0.0001 | 465724 | 101286 | 1.24 (1.23-1.25) | < 0.0001 | 465724 | 101286 | 1.25 (1.25-1.26) | < 0.0001 |
| 70 | **Poisoning** | 493033 | 3722 | 1.19 (1.15-1.22) | < 0.0001 | 493033 | 3722 | 1.24 (1.20-1.27) | < 0.0001 | 493033 | 3722 | 1.27 (1.23-1.31) | < 0.0001 |
| 13 | **Blood diseases** | 484225 | 52131 | 1.18 (1.17-1.19) | < 0.0001 | 484225 | 52131 | 1.22 (1.21-1.23) | < 0.0001 | 484225 | 52131 | 1.23 (1.22-1.24) | < 0.0001 |
| 43 | *Cerebral infarction* | 494133 | 10211 | 1.18 (1.16-1.21) | < 0.0001 | 494133 | 10211 | 1.21 (1.18-1.23) | < 0.0001 | 494133 | 10211 | 1.23 (1.20-1.25) | < 0.0001 |
| 17 | **Mental and behavioural disorders** | 486370 | 76820 | 1.17 (1.16-1.18) | < 0.0001 | 486370 | 76820 | 1.21 (1.20-1.22) | < 0.0001 | 486370 | 76820 | 1.23 (1.22-1.24) | < 0.0001 |
| 0 | **All-cause mortality** | 495900 | 42753 | 1.16 (1.15-1.18) | < 0.0001 | 495900 | 42753 | 1.22 (1.21-1.23) | < 0.0001 | 495900 | 42753 | 1.23 (1.22-1.24) | < 0.0001 |
| 48 | Chronic obstructive pulmonary disease | 493661 | 23358 | 1.16 (1.15-1.18) | < 0.0001 | 493661 | 23358 | 1.27 (1.25-1.28) | < 0.0001 | 493661 | 23358 | 1.30 (1.29-1.32) | < 0.0001 |
| 22 | Neurotic disorders | 493541 | 26384 | 1.15 (1.14-1.16) | < 0.0001 | 493541 | 26384 | 1.20 (1.18-1.21) | < 0.0001 | 493541 | 26384 | 1.20 (1.19-1.22) | < 0.0001 |
| 59 | Rheumatoid arthritis | 485066 | 62314 | 1.15 (1.14-1.16) | < 0.0001 | 485066 | 62314 | 1.17 (1.16-1.18) | < 0.0001 | 485066 | 62314 | 1.19 (1.18-1.19) | < 0.0001 |
| 20 | Psychotic disorders | 495210 | 1585 | 1.15 (1.10-1.20) | < 0.0001 | 495210 | 1585 | 1.26 (1.21-1.32) | < 0.0001 | 495210 | 1585 | 1.28 (1.23-1.34) | < 0.0001 |
| 40 | Cerebrovascular diseases | 492411 | 22282 | 1.14 (1.13-1.15) | < 0.0001 | 492411 | 22282 | 1.17 (1.16-1.19) | < 0.0001 | 492411 | 22282 | 1.20 (1.18-1.21) | < 0.0001 |
| 41 | Stroke | 493269 | 13313 | 1.13 (1.12-1.15) | < 0.0001 | 493269 | 13313 | 1.16 (1.14-1.18) | < 0.0001 | 493269 | 13313 | 1.17 (1.15-1.19) | < 0.0001 |
| 50 | **Digestive system diseases** | 406095 | 174068 | 1.13 (1.12-1.13) | < 0.0001 | 406095 | 174068 | 1.15 (1.15-1.16) | < 0.0001 | 406095 | 174068 | 1.16 (1.15-1.17) | < 0.0001 |
| 19 | Substance use disorders | 492516 | 30983 | 1.12 (1.11-1.13) | < 0.0001 | 492516 | 30983 | 1.16 (1.15-1.17) | < 0.0001 | 492516 | 30983 | 1.18 (1.17-1.20) | < 0.0001 |
| 28 | Transient ischaemic attack | 494579 | 4802 | 1.12 (1.09-1.15) | < 0.0001 | 494579 | 4802 | 1.14 (1.11-1.18) | < 0.0001 | 494579 | 4802 | 1.16 (1.12-1.19) | < 0.0001 |
| 73 | **Self-harm** | 493783 | 2087 | 1.12 (1.08-1.17) | < 0.0001 | 493783 | 2087 | 1.18 (1.13-1.23) | < 0.0001 | 493783 | 2087 | 1.22 (1.17-1.27) | < 0.0001 |
| 26 | Epilepsy | 494157 | 5544 | 1.11 (1.08-1.13) | < 0.0001 | 494157 | 5544 | 1.14 (1.11-1.17) | < 0.0001 | 494157 | 5544 | 1.17 (1.14-1.20) | < 0.0001 |
| 30 | **Eye diseases** | 476556 | 84729 | 1.10 (1.09-1.11) | < 0.0001 | 476556 | 84729 | 1.12 (1.12-1.13) | < 0.0001 | 476556 | 84729 | 1.13 (1.13-1.14) | < 0.0001 |
| 52 | Inflammatory bowel disease | 484041 | 19742 | 1.09 (1.08-1.11) | < 0.0001 | 484041 | 19742 | 1.13 (1.11-1.14) | < 0.0001 | 484041 | 19742 | 1.14 (1.13-1.16) | < 0.0001 |
| 5 | Colorectal cancer | 493983 | 8227 | 1.09 (1.07-1.11) | < 0.0001 | 493983 | 8227 | 1.13 (1.10-1.15) | < 0.0001 | 493983 | 8227 | 1.11 (1.08-1.13) | < 0.0001 |
| 68 | **Digestive and abdominal symptoms** | 460697 | 79339 | 1.08 (1.08-1.09) | < 0.0001 | 460697 | 79339 | 1.11 (1.10-1.12) | < 0.0001 | 460697 | 79339 | 1.12 (1.11-1.13) | < 0.0001 |
| 3 | Viral infections | 492884 | 6574 | 1.08 (1.06-1.11) | < 0.0001 | 492884 | 6574 | 1.10 (1.07-1.12) | < 0.0001 | 492884 | 6574 | 1.11 (1.08-1.13) | < 0.0001 |
| 31 | **Ear diseases** | 493777 | 3410 | 1.08 (1.04-1.11) | 0.0003 | 493777 | 3410 | 1.10 (1.07-1.14) | < 0.0001 | 493777 | 3410 | 1.11 (1.08-1.15) | < 0.0001 |
| 72 | **Fall** | 485173 | 36667 | 1.07 (1.06-1.09) | < 0.0001 | 485173 | 36667 | 1.12 (1.11-1.14) | < 0.0001 | 485173 | 36667 | 1.12 (1.11-1.13) | < 0.0001 |
| 27 | Headaches | 494187 | 7391 | 1.06 (1.04-1.09) | < 0.0001 | 494187 | 7391 | 1.07 (1.05-1.10) | < 0.0001 | 494187 | 7391 | 1.06 (1.04-1.09) | < 0.0001 |
| 8 | Breast cancer (Female) | 262868 | 12224 | 1.06 (1.04-1.08) | < 0.0001 | 262868 | 12224 | 1.08 (1.06-1.10) | < 0.0001 | 262868 | 12224 | 1.05 (1.03-1.07) | < 0.0001 |
| 18 | Dementia | 495824 | 9244 | 1.05 (1.03-1.07) | 0.0005 | 495824 | 9244 | 1.08 (1.05-1.10) | < 0.0001 | 495824 | 9244 | 1.12 (1.10-1.15) | < 0.0001 |
| 51 | Appendicitis | 493676 | 2753 | 1.05 (1.01-1.09) | 0.09 | 493676 | 2753 | 1.06 (1.02-1.10) | 0.04 | 493676 | 2753 | 1.04 (1.00-1.08) | 0.44 |
| 69 | **Injury** | 471296 | 53225 | 1.04 (1.03-1.05) | < 0.0001 | 471296 | 53225 | 1.08 (1.07-1.09) | < 0.0001 | 471296 | 53225 | 1.07 (1.06-1.08) | < 0.0001 |
| 12 | Leukaemia / Lymphoma | 494411 | 12409 | 1.04 (1.02-1.06) | 0.0001 | 494411 | 12409 | 1.08 (1.06-1.10) | < 0.0001 | 494411 | 12409 | 1.05 (1.03-1.07) | < 0.0001 |
| 4 | **Cancers** | 475215 | 80417 | 1.03 (1.03-1.04) | < 0.0001 | 475215 | 80417 | 1.06 (1.05-1.06) | < 0.0001 | 475215 | 80417 | 1.03 (1.03-1.04) | < 0.0001 |
| 24 | Parkinson disease | 495694 | 4058 | 1.02 (0.98-1.05) | 1.00 | 495694 | 4058 | 1.07 (1.03-1.10) | 0.002 | 495694 | 4058 | 1.06 (1.03-1.09) | 0.005 |
| 42 | *Intracerebral haemorrhage* | 494971 | 3905 | 1.01 (0.97-1.04) | 1.00 | 494971 | 3905 | 1.03 (1.00-1.07) | 0.46 | 494971 | 3905 | 1.04 (1.00-1.07) | 0.30 |
| 6 | Lung cancer | 495622 | 6298 | 0.98 (0.95-1.00) | 0.64 | 495622 | 6298 | 1.05 (1.03-1.08) | 0.002 | 495622 | 6298 | 1.05 (1.03-1.08) | 0.002 |
| 11 | Brain cancer | 495805 | 1287 | 0.98 (0.93-1.04) | 1.00 | 495805 | 1287 | 0.98 (0.93-1.04) | 1.00 | 495805 | 1287 | 0.97 (0.91-1.02) | 1.00 |
| 9 | Prostate cancer (Male) | 224487 | 13355 | 0.96 (0.94-0.98) | < 0.0001 | 224487 | 13355 | 0.98 (0.96-0.99) | 0.010 | 224487 | 13355 | 0.96 (0.95-0.98) | < 0.0001 |
| 7 | Melanoma | 490441 | 24183 | 0.94 (0.92-0.95) | < 0.0001 | 490441 | 24183 | 0.95 (0.94-0.97) | < 0.0001 | 490441 | 24183 | 0.92 (0.91-0.94) | < 0.0001 |
| 25 | Multiple sclerosis | 495244 | 1341 | 0.93 (0.88-0.98) | 0.12 | 495244 | 1341 | 1.02 (0.96-1.07) | 1.00 | 495244 | 1341 | 1.00 (0.95-1.06) | 1.00 |
| 71 | **Road accidents** | 491968 | 5978 | 0.90 (0.87-0.92) | < 0.0001 | 491968 | 5978 | 0.90 (0.88-0.92) | < 0.0001 | 491968 | 5978 | 0.87 (0.84-0.89) | < 0.0001 |

Supplement eTable 5. Difference in population attributable fractions between obesity defined using different measures

| | | $PAF_{WC}$ - $PAF_{BMI}$ | | | $PAF_{WHtR}$ - $PAF_{BMI}$ | | |
|---|---|---|---|---|---|---|---|
| | | % | Lower CI | Upper CI | % | Lower CI | Upper CI |
| 0 | **All-cause mortality** | 0.18 | -1.97 | 2.34 | 0.19 | -1.97 | 2.35 |
| 0.1 | **Simple multimorbidity (2nd disease)** | 0.06 | -0.76 | 0.87 | 0.07 | -0.75 | 0.88 |
| 0.2 | **Third disease** | 0.14 | -0.85 | 1.13 | 0.15 | -0.84 | 1.14 |
| 0.3 | **Complex multimorbidity (4th disease)** | 0.17 | -1.00 | 1.35 | 0.19 | -0.99 | 1.37 |
| 1 | **Infectious diseases** | 0.14 | -1.46 | 1.74 | 0.15 | -1.45 | 1.75 |
| 2 | Bacterial infections | 0.11 | -1.95 | 2.17 | 0.12 | -1.94 | 2.18 |
| 3 | Viral infections | 0.59 | -4.65 | 5.83 | 0.58 | -4.66 | 5.82 |
| 4 | **Cancers** | 0.10 | -1.39 | 1.59 | 0.10 | -1.39 | 1.59 |
| 5 | Colorectal cancer | 0.15 | -4.99 | 5.29 | 0.14 | -5.00 | 5.28 |
| 6 | Lung cancer | 0.04 | -6.05 | 6.12 | 0.03 | -6.05 | 6.12 |
| 7 | Melanoma | -0.01 | -2.94 | 2.93 | -0.01 | -2.94 | 2.92 |
| 8 | Breast cancer (Female) | 0.06 | -3.06 | 3.19 | 0.06 | -3.06 | 3.19 |
| 9 | Prostate cancer (Male) | -0.04 | -5.35 | 5.27 | -0.11 | -5.42 | 5.20 |
| 10 | Kidney cancer | -0.31 | -9.37 | 8.74 | -0.32 | -9.37 | 8.74 |
| 11 | Brain cancer | -0.11 | -14.53 | 14.31 | -0.11 | -14.53 | 14.31 |
| 12 | Leukaemia / Lymphoma | 0.20 | -3.95 | 4.36 | 0.20 | -3.96 | 4.36 |
| 13 | **Blood diseases** | 0.03 | -1.73 | 1.79 | 0.05 | -1.72 | 1.81 |
| 14 | Anaemia | 0.03 | -1.85 | 1.91 | 0.05 | -1.83 | 1.93 |
| 15 | **Endocrine diseases** | 0.08 | -0.78 | 0.95 | 0.09 | -0.77 | 0.96 |
| 16 | Diabetes mellitus | 0.19 | -0.90 | 1.28 | 0.19 | -0.90 | 1.28 |
| 17 | **Mental and behavioural disorders** | 0.06 | -1.32 | 1.43 | 0.07 | -1.31 | 1.44 |
| 18 | Dementia | 0.08 | -5.02 | 5.17 | 0.07 | -5.02 | 5.17 |
| 19 | Substance use disorders | 0.11 | -2.03 | 2.26 | 0.11 | -2.03 | 2.25 |
| 20 | Psychotic disorders | 0.52 | -9.67 | 10.70 | 0.51 | -9.67 | 10.70 |
| 21 | Mood disorders | 0.16 | -1.99 | 2.32 | 0.18 | -1.98 | 2.33 |
| 22 | Neurotic disorders | 0.17 | -2.24 | 2.59 | 0.17 | -2.25 | 2.59 |
| 23 | **Neurological disorders** | 0.05 | -1.41 | 1.52 | 0.08 | -1.38 | 1.55 |
| 24 | Parkinson disease | 0.00 | -8.02 | 8.02 | 0.00 | -8.02 | 8.02 |
| 25 | Multiple sclerosis | 0.25 | -10.16 | 10.66 | 0.25 | -10.16 | 10.66 |
| 26 | Epilepsy | -0.17 | -6.03 | 5.70 | -0.17 | -6.04 | 5.69 |
| 27 | Headaches | 0.41 | -4.14 | 4.96 | 0.40 | -4.15 | 4.95 |
| 28 | Transient ischaemic attack | -0.27 | -6.93 | 6.40 | -0.27 | -6.93 | 6.40 |
| 29 | Sleep disorders | -0.03 | -2.23 | 2.17 | 0.00 | -2.20 | 2.20 |
| 30 | **Eye diseases** | 0.01 | -1.38 | 1.40 | 0.02 | -1.37 | 1.41 |
| 31 | **Ear diseases** | 0.37 | -7.02 | 7.77 | 0.52 | -6.87 | 7.91 |
| 32 | **Circulatory diseases** | 0.03 | -0.75 | 0.82 | 0.03 | -0.75 | 0.82 |
| 33 | Hypertension | 0.03 | -0.89 | 0.96 | 0.04 | -0.89 | 0.96 |
| 34 | Ischaemic heart disease | 0.18 | -1.65 | 2.02 | 0.18 | -1.65 | 2.02 |
| 35 | *Angina pectoris* | 0.21 | -2.36 | 2.78 | 0.21 | -2.37 | 2.78 |
| 36 | *Myocardial infarction* | 0.12 | -3.67 | 3.91 | 0.11 | -3.68 | 3.91 |
| 37 | Pulmonary embolism | 0.19 | -3.93 | 4.31 | 0.19 | -3.93 | 4.30 |
| 38 | Arrhythmias | 0.07 | -1.90 | 2.04 | 0.05 | -1.92 | 2.03 |
| 39 | Heart failure | -0.08 | -2.75 | 2.58 | -0.09 | -2.75 | 2.58 |
| 40 | Cerebrovascular diseases | 0.07 | -3.01 | 3.15 | 0.06 | -3.02 | 3.14 |
| 41 | Stroke | 0.24 | -3.80 | 4.28 | 0.24 | -3.80 | 4.28 |
| 42 | *Intracerebral haemorrhage* | 0.41 | -7.23 | 8.05 | 0.41 | -7.24 | 8.05 |
| 43 | *Cerebral infarction* | 0.20 | -4.37 | 4.77 | 0.20 | -4.38 | 4.77 |
| 44 | Arteriosclerosis | 0.48 | -7.14 | 8.10 | 0.48 | -7.15 | 8.10 |
| 45 | Deep vein thrombosis | 0.24 | -4.34 | 4.82 | 0.24 | -4.34 | 4.82 |
| 46 | **Respiratory diseases** | 0.16 | -1.04 | 1.36 | 0.17 | -1.03 | 1.36 |
| 47 | Respiratory tract infections | 0.14 | -2.23 | 2.52 | 0.19 | -2.18 | 2.56 |
| 48 | Chronic obstructive pulmonary disease | 0.44 | -2.36 | 3.23 | 0.44 | -2.36 | 3.23 |
| 49 | Asthma | 0.24 | -1.66 | 2.15 | 0.24 | -1.66 | 2.15 |
| 50 | **Digestive system diseases** | 0.06 | -0.73 | 0.86 | 0.07 | -0.72 | 0.86 |
| 51 | Appendicitis | -0.03 | -7.58 | 7.53 | -0.03 | -7.59 | 7.52 |
| 52 | Inflammatory bowel disease | 0.28 | -2.68 | 3.24 | 0.30 | -2.66 | 3.26 |

| | | | | | | | |
|---|---|---|---|---|---|---|---|
| 53 | Liver diseases | 0.21 | -2.57 | 2.99 | 0.24 | -2.54 | 3.01 |
| 54 | Alcohol-related liver diseases | 0.60 | -8.86 | 10.07 | 0.60 | -8.87 | 10.07 |
| 55 | Pancreatitis | -0.36 | -7.00 | 6.28 | -0.36 | -7.00 | 6.28 |
| 56 | **Skin diseases** | 0.19 | -1.43 | 1.82 | 0.20 | -1.43 | 1.83 |
| 57 | Eczema and skin infections | 0.38 | -2.00 | 2.76 | 0.37 | -2.01 | 2.76 |
| 58 | **Musculoskeletal diseases** | 0.01 | -0.85 | 0.87 | 0.02 | -0.85 | 0.88 |
| 59 | Rheumatoid arthritis | 0.21 | -1.33 | 1.74 | 0.22 | -1.32 | 1.76 |
| 60 | Gout | -0.06 | -2.95 | 2.83 | -0.06 | -2.95 | 2.83 |
| 61 | Osteoarthritis | -0.02 | -1.23 | 1.19 | -0.01 | -1.22 | 1.21 |
| 62 | Sciatica | -0.05 | -3.23 | 3.13 | 0.01 | -3.17 | 3.19 |
| 63 | Backpain | 0.03 | -2.35 | 2.41 | 0.08 | -2.30 | 2.46 |
| 64 | Soft tissue diseases | 0.01 | -1.71 | 1.73 | 0.03 | -1.70 | 1.75 |
| 65 | **Genitourinary diseases** | -0.01 | -1.16 | 1.13 | -0.01 | -1.16 | 1.13 |
| 66 | Renal failure | 0.10 | -1.75 | 1.96 | 0.12 | -1.73 | 1.98 |
| 67 | **Circulatory and respiratory symptoms** | 0.07 | -1.40 | 1.54 | 0.07 | -1.40 | 1.54 |
| 68 | **Digestive and abdominal symptoms** | 0.03 | -1.32 | 1.39 | 0.05 | -1.31 | 1.40 |
| 69 | **Injury** | 0.15 | -1.62 | 1.91 | 0.17 | -1.60 | 1.94 |
| 70 | **Poisoning** | 0.69 | -5.86 | 7.25 | 0.69 | -5.86 | 7.25 |
| 71 | **Road accidents** | 0.24 | -4.99 | 5.48 | 0.24 | -5.00 | 5.48 |
| 72 | **Fall** | 0.15 | -2.02 | 2.31 | 0.17 | -2.00 | 2.34 |
| 73 | **Self-harm** | 0.19 | -8.34 | 8.72 | 0.19 | -8.34 | 8.72 |

Supplement eTable 6. Difference in population attributable fractions between obesity class defined using different measures

| | | BMI 30-34 and equivalents | | | | | | BMI ≥35 and equivalents | | | | | |
|---|---|---|---|---|---|---|---|---|---|---|---|---|---|
| | | $PAF_{WC}$ - $PAF_{BMI}$ | | | $PAF_{WHtR}$ - $PAF_{BMI}$ | | | $PAF_{WC}$ - $PAF_{BMI}$ | | | $PAF_{WHtR}$ - $PAF_{BMI}$ | | |
| | | % | Lower CI | Upper CI | % | Lower CI | Upper CI | % | Lower CI | Upper CI | % | Lower CI | Upper CI |
| 0 | **All-cause mortality** | 0.25 | -1.63 | 2.14 | 0.27 | -1.62 | 2.15 | 0.00 | -1.57 | 1.56 | 0.00 | -1.57 | 1.56 |
| 0.1 | **Simple multimorbidity (2nd disease)** | 0.10 | -0.71 | 0.91 | 0.11 | -0.70 | 0.92 | 0.00 | -0.75 | 0.76 | 0.00 | -0.75 | 0.76 |
| 0.2 | **Third disease** | 0.26 | -0.86 | 1.38 | 0.27 | -0.85 | 1.39 | 0.00 | -1.05 | 1.05 | 0.00 | -1.05 | 1.05 |
| 0.3 | **Complex multimorbidity (4th disease)** | 0.36 | -1.13 | 1.84 | 0.37 | -1.12 | 1.85 | 0.00 | -1.33 | 1.33 | 0.00 | -1.33 | 1.33 |
| 1 | **Infectious diseases** | 0.25 | -1.17 | 1.67 | 0.27 | -1.15 | 1.69 | 0.00 | -1.21 | 1.21 | 0.00 | -1.21 | 1.21 |
| 2 | Bacterial infections | 0.30 | -1.57 | 2.16 | 0.33 | -1.54 | 2.19 | 0.00 | -1.60 | 1.60 | 0.00 | -1.60 | 1.60 |
| 3 | Viral infections | 0.38 | -3.86 | 4.62 | 0.38 | -3.86 | 4.63 | 0.00 | -3.63 | 3.63 | 0.00 | -3.63 | 3.63 |
| 4 | **Cancers** | 0.07 | -1.09 | 1.23 | 0.06 | -1.10 | 1.22 | 0.00 | -0.91 | 0.91 | 0.00 | -0.91 | 0.91 |
| 5 | Colorectal cancer | -0.01 | -4.23 | 4.20 | -0.01 | -4.22 | 4.21 | 0.00 | -3.43 | 3.43 | 0.00 | -3.43 | 3.43 |
| 6 | Lung cancer | -0.04 | -4.79 | 4.71 | -0.04 | -4.78 | 4.71 | 0.00 | -3.80 | 3.80 | 0.00 | -3.80 | 3.80 |
| 7 | Melanoma | 0.00 | -2.12 | 2.11 | -0.03 | -2.14 | 2.09 | 0.00 | -1.52 | 1.52 | 0.00 | -1.52 | 1.52 |
| 8 | Breast cancer (Female) | 0.15 | -2.26 | 2.55 | 0.13 | -2.28 | 2.53 | 0.00 | -2.13 | 2.13 | 0.00 | -2.13 | 2.13 |
| 9 | Prostate cancer (Male) | 0.05 | -3.82 | 3.91 | -0.02 | -3.88 | 3.85 | 0.00 | -2.63 | 2.63 | 0.00 | -2.63 | 2.63 |
| 10 | Kidney cancer | -0.43 | -9.56 | 8.69 | -0.43 | -9.56 | 8.69 | 0.00 | -7.80 | 7.80 | 0.00 | -7.80 | 7.80 |
| 11 | Brain cancer | -0.21 | -10.72 | 10.30 | -0.21 | -10.72 | 10.30 | 0.00 | -7.65 | 7.65 | 0.00 | -7.65 | 7.65 |
| 12 | Leukaemia / Lymphoma | 0.17 | -3.11 | 3.45 | 0.13 | -3.15 | 3.41 | 0.00 | -2.60 | 2.59 | 0.00 | -2.60 | 2.59 |
| 13 | **Blood diseases** | 0.12 | -1.44 | 1.67 | 0.13 | -1.43 | 1.69 | 0.00 | -1.32 | 1.32 | 0.00 | -1.32 | 1.32 |
| 14 | Anaemia | 0.10 | -1.58 | 1.79 | 0.12 | -1.57 | 1.81 | 0.00 | -1.44 | 1.44 | 0.00 | -1.44 | 1.44 |
| 15 | **Endocrine diseases** | 0.13 | -0.80 | 1.06 | 0.14 | -0.79 | 1.08 | 0.00 | -0.89 | 0.89 | 0.00 | -0.89 | 0.89 |
| 16 | Diabetes mellitus | 0.38 | -1.50 | 2.26 | 0.38 | -1.50 | 2.26 | 0.00 | -1.50 | 1.50 | 0.00 | -1.50 | 1.50 |
| 17 | **Mental and behavioural disorders** | 0.06 | -1.13 | 1.25 | 0.07 | -1.12 | 1.26 | 0.00 | -1.02 | 1.02 | 0.00 | -1.02 | 1.02 |
| 18 | Dementia | 0.20 | -3.92 | 4.32 | 0.21 | -3.91 | 4.33 | 0.00 | -3.30 | 3.30 | 0.00 | -3.30 | 3.30 |
| 19 | Substance use disorders | 0.04 | -1.73 | 1.81 | 0.04 | -1.73 | 1.81 | 0.00 | -1.48 | 1.48 | 0.00 | -1.48 | 1.48 |
| 20 | Psychotic disorders | 0.78 | -8.18 | 9.75 | 0.78 | -8.18 | 9.75 | 0.00 | -7.87 | 7.87 | 0.00 | -7.87 | 7.87 |
| 21 | Mood disorders | 0.14 | -1.92 | 2.21 | 0.16 | -1.90 | 2.23 | 0.00 | -1.83 | 1.83 | 0.00 | -1.83 | 1.83 |
| 22 | Neurotic disorders | 0.24 | -1.82 | 2.31 | 0.25 | -1.82 | 2.31 | 0.00 | -1.81 | 1.81 | 0.00 | -1.81 | 1.81 |
| 23 | **Neurological disorders** | 0.03 | -1.32 | 1.38 | 0.08 | -1.27 | 1.43 | 0.00 | -1.17 | 1.17 | 0.00 | -1.17 | 1.17 |
| 24 | Parkinson disease | -0.17 | -6.50 | 6.17 | -0.16 | -6.50 | 6.17 | 0.00 | -4.91 | 4.91 | 0.00 | -4.91 | 4.91 |
| 25 | Multiple sclerosis | 0.40 | -6.93 | 7.72 | 0.40 | -6.92 | 7.72 | 0.00 | -6.00 | 5.99 | 0.00 | -6.00 | 5.99 |
| 26 | Epilepsy | 0.09 | -4.85 | 5.03 | 0.09 | -4.85 | 5.03 | 0.00 | -4.15 | 4.15 | 0.00 | -4.15 | 4.15 |
| 27 | Headaches | 0.34 | -3.14 | 3.83 | 0.35 | -3.13 | 3.83 | 0.00 | -3.08 | 3.08 | 0.00 | -3.08 | 3.08 |
| 28 | Transient ischaemic attack | -0.56 | -6.30 | 5.18 | -0.56 | -6.29 | 5.18 | 0.00 | -4.83 | 4.82 | 0.00 | -4.83 | 4.82 |
| 29 | Sleep disorders | -0.15 | -4.27 | 3.98 | -0.07 | -4.20 | 4.05 | 0.00 | -2.51 | 2.51 | 0.00 | -2.51 | 2.51 |
| 30 | **Eye diseases** | -0.01 | -1.16 | 1.13 | 0.01 | -1.14 | 1.15 | 0.01 | -0.94 | 0.96 | 0.01 | -0.94 | 0.96 |
| 31 | **Ear diseases** | 0.70 | -5.34 | 6.74 | 0.85 | -5.18 | 6.89 | 0.00 | -4.92 | 4.92 | 0.00 | -4.92 | 4.92 |
| 32 | **Circulatory diseases** | 0.07 | -0.65 | 0.80 | 0.08 | -0.65 | 0.80 | 0.00 | -0.65 | 0.65 | 0.00 | -0.65 | 0.65 |
| 33 | Hypertension | 0.12 | -0.88 | 1.11 | 0.13 | -0.86 | 1.13 | 0.00 | -0.95 | 0.95 | 0.00 | -0.95 | 0.95 |
| 34 | Ischaemic heart disease | 0.36 | -1.48 | 2.19 | 0.36 | -1.48 | 2.19 | 0.00 | -1.63 | 1.63 | 0.00 | -1.63 | 1.63 |
| 35 | *Angina pectoris* | 0.36 | -2.37 | 3.09 | 0.36 | -2.37 | 3.09 | 0.00 | -2.49 | 2.48 | 0.00 | -2.49 | 2.48 |
| 36 | *Myocardial infarction* | 0.34 | -3.24 | 3.92 | 0.34 | -3.24 | 3.92 | 0.00 | -3.17 | 3.17 | 0.00 | -3.17 | 3.17 |
| 37 | Pulmonary embolism | 0.18 | -4.24 | 4.61 | 0.11 | -4.31 | 4.54 | 0.00 | -3.81 | 3.81 | 0.00 | -3.81 | 3.81 |
| 38 | Arrhythmias | 0.04 | -1.82 | 1.90 | 0.00 | -1.86 | 1.86 | 0.00 | -1.57 | 1.57 | 0.00 | -1.57 | 1.57 |
| 39 | Heart failure | -0.19 | -3.36 | 2.98 | -0.19 | -3.36 | 2.99 | 0.00 | -2.55 | 2.55 | 0.00 | -2.55 | 2.55 |
| 40 | Cerebrovascular diseases | 0.10 | -2.58 | 2.77 | 0.10 | -2.57 | 2.77 | 0.00 | -2.24 | 2.24 | 0.00 | -2.24 | 2.24 |
| 41 | Stroke | 0.23 | -3.21 | 3.66 | 0.23 | -3.21 | 3.66 | 0.00 | -2.83 | 2.83 | 0.00 | -2.83 | 2.83 |
| 42 | *Intracerebral haemorrhage* | 0.33 | -5.40 | 6.07 | 0.33 | -5.40 | 6.07 | 0.00 | -4.57 | 4.57 | 0.00 | -4.57 | 4.57 |
| 43 | *Cerebral infarction* | 0.13 | -3.98 | 4.24 | 0.13 | -3.98 | 4.24 | 0.00 | -3.42 | 3.42 | 0.00 | -3.42 | 3.42 |
| 44 | Arteriosclerosis | 0.41 | -6.73 | 7.54 | 0.41 | -6.73 | 7.54 | 0.00 | -6.07 | 6.07 | 0.00 | -6.07 | 6.07 |
| 45 | Deep vein thrombosis | 0.61 | -3.86 | 5.09 | 0.62 | -3.86 | 5.09 | 0.00 | -3.90 | 3.90 | 0.00 | -3.90 | 3.90 |
| 46 | **Respiratory diseases** | 0.21 | -0.84 | 1.27 | 0.22 | -0.84 | 1.27 | 0.00 | -0.89 | 0.89 | 0.00 | -0.89 | 0.89 |
| 47 | Respiratory tract infections | 0.24 | -2.00 | 2.49 | 0.30 | -1.94 | 2.54 | 0.00 | -1.90 | 1.90 | 0.00 | -1.90 | 1.90 |
| 48 | Chronic obstructive pulmonary disease | 0.76 | -1.78 | 3.31 | 0.79 | -1.76 | 3.33 | 0.00 | -2.18 | 2.18 | 0.00 | -2.18 | 2.18 |
| 49 | Asthma | 0.30 | -1.53 | 2.12 | 0.30 | -1.52 | 2.13 | 0.01 | -1.58 | 1.61 | 0.01 | -1.58 | 1.61 |
| 50 | **Digestive system diseases** | 0.11 | -0.54 | 0.75 | 0.11 | -0.53 | 0.75 | 0.00 | -0.51 | 0.52 | 0.00 | -0.51 | 0.52 |
| 51 | Appendicitis | -0.11 | -5.97 | 5.74 | -0.11 | -5.96 | 5.74 | 0.00 | -4.63 | 4.63 | 0.00 | -4.63 | 4.63 |
| 52 | Inflammatory bowel disease | 0.33 | -2.09 | 2.76 | 0.36 | -2.06 | 2.79 | 0.00 | -2.04 | 2.04 | 0.00 | -2.04 | 2.04 |
| 53 | Liver diseases | 0.40 | -2.87 | 3.67 | 0.40 | -2.87 | 3.67 | 0.00 | -2.84 | 2.84 | 0.00 | -2.84 | 2.84 |
| 54 | Alcohol-related liver diseases | 1.06 | -8.77 | 10.89 | 1.06 | -8.77 | 10.89 | 0.00 | -8.80 | 8.80 | 0.00 | -8.80 | 8.80 |
| 55 | Pancreatitis | -0.38 | -7.67 | 6.92 | -0.38 | -7.67 | 6.92 | 0.00 | -6.58 | 6.58 | 0.00 | -6.58 | 6.58 |
| 56 | **Skin diseases** | 0.18 | -1.23 | 1.60 | 0.18 | -1.23 | 1.60 | 0.00 | -1.19 | 1.19 | 0.00 | -1.19 | 1.19 |
| 57 | Eczema and skin infections | 0.40 | -2.09 | 2.90 | 0.38 | -2.12 | 2.88 | 0.00 | -2.08 | 2.08 | 0.00 | -2.08 | 2.08 |
| 58 | **Musculoskeletal diseases** | 0.01 | -0.75 | 0.76 | 0.02 | -0.73 | 0.78 | 0.00 | -0.64 | 0.64 | 0.00 | -0.64 | 0.64 |
| 59 | Rheumatoid arthritis | 0.27 | -1.02 | 1.57 | 0.30 | -1.00 | 1.59 | 0.01 | -1.11 | 1.12 | 0.01 | -1.11 | 1.12 |
| 60 | Gout | -0.23 | -4.43 | 3.97 | -0.23 | -4.44 | 3.97 | 0.00 | -3.72 | 3.72 | 0.00 | -3.72 | 3.72 |
| 61 | Osteoarthritis | 0.00 | -1.23 | 1.23 | 0.02 | -1.21 | 1.25 | 0.01 | -1.11 | 1.13 | 0.01 | -1.11 | 1.13 |
| 62 | Sciatica | -0.21 | -3.17 | 2.75 | -0.14 | -3.10 | 2.82 | 0.00 | -2.62 | 2.62 | 0.00 | -2.62 | 2.62 |
| 63 | Backpain | 0.14 | -2.10 | 2.38 | 0.16 | -2.08 | 2.40 | 0.00 | -1.98 | 1.98 | 0.00 | -1.98 | 1.98 |
| 64 | Soft tissue diseases | 0.07 | -1.50 | 1.64 | 0.10 | -1.48 | 1.67 | 0.00 | -1.34 | 1.34 | 0.00 | -1.34 | 1.34 |
| 65 | **Genitourinary diseases** | -0.01 | -1.00 | 0.98 | 0.00 | -0.99 | 0.99 | 0.00 | -0.84 | 0.83 | 0.00 | -0.84 | 0.83 |
| 66 | Renal failure | 0.24 | -1.84 | 2.31 | 0.27 | -1.81 | 2.35 | 0.00 | -1.81 | 1.81 | 0.00 | -1.81 | 1.81 |
| 67 | **Circulatory and respiratory symptoms** | 0.08 | -1.23 | 1.38 | 0.09 | -1.22 | 1.39 | 0.00 | -1.12 | 1.12 | 0.00 | -1.12 | 1.12 |
| 68 | **Digestive and abdominal symptoms** | 0.10 | -0.99 | 1.19 | 0.11 | -0.98 | 1.20 | 0.00 | -0.90 | 0.89 | 0.00 | -0.90 | 0.89 |
| 69 | **Injury** | 0.15 | -1.23 | 1.53 | 0.16 | -1.21 | 1.54 | 0.00 | -1.11 | 1.11 | 0.00 | -1.11 | 1.11 |
| 70 | **Poisoning** | 0.93 | -4.83 | 6.69 | 0.93 | -4.83 | 6.69 | 0.00 | -5.00 | 5.00 | 0.00 | -5.00 | 5.00 |
| 71 | **Road accidents** | 0.25 | -3.28 | 3.79 | 0.18 | -3.35 | 3.71 | 0.00 | -2.55 | 2.56 | 0.00 | -2.55 | 2.56 |
| 72 | **Fall** | 0.15 | -1.59 | 1.90 | 0.18 | -1.56 | 1.93 | 0.00 | -1.45 | 1.45 | 0.00 | -1.45 | 1.45 |
| 73 | **Self-harm** | 0.12 | -6.88 | 7.11 | 0.12 | -6.87 | 7.11 | 0.00 | -6.00 | 6.00 | 0.00 | -6.00 | 6.00 |

Supplement eTable 7. Difference in population attributable fractions between overweight defined using different measures

| | | $PAF_{WC}$ - $PAF_{BMI}$ | | | $PAF_{WHtR}$ - $PAF_{BMI}$ | | |
|---|---|---|---|---|---|---|---|
| | | % | Lower CI | Upper CI | % | Lower CI | Upper CI |
| 0 | **All-cause mortality** | 5.35 | 3.28 | 7.42 | 7.17 | 5.13 | 9.22 |
| 0.1 | **Simple multimorbidity (2nd disease)** | 2.13 | 1.27 | 3.00 | 2.84 | 1.99 | 3.69 |
| 0.2 | **Third disease** | 4.09 | 2.91 | 5.28 | 5.04 | 3.87 | 6.21 |
| 0.3 | **Complex multimorbidity (4th disease)** | 5.76 | 4.13 | 7.38 | 7.16 | 5.56 | 8.77 |
| 1 | **Infectious diseases** | 4.75 | 3.18 | 6.32 | 5.55 | 4.00 | 7.09 |
| 2 | Bacterial infections | 6.43 | 4.39 | 8.47 | 7.27 | 5.27 | 9.28 |
| 3 | Viral infections | -0.76 | -5.52 | 4.01 | 0.61 | -4.06 | 5.27 |
| 4 | **Cancers** | 2.40 | 1.10 | 3.70 | 0.24 | -1.04 | 1.52 |
| 5 | Colorectal cancer | 3.84 | -0.69 | 8.37 | 1.63 | -2.84 | 6.11 |
| 6 | Lung cancer | 7.10 | 1.97 | 12.23 | 7.34 | 2.23 | 12.45 |
| 7 | Melanoma | 1.36 | -1.10 | 3.82 | -1.77 | -4.19 | 0.65 |
| 8 | Breast cancer (Female) | 3.21 | 0.44 | 5.97 | 0.86 | -1.91 | 3.63 |
| 9 | Prostate cancer (Male) | 0.63 | -3.39 | 4.64 | -1.75 | -5.61 | 2.11 |
| 10 | Kidney cancer | 5.95 | -3.20 | 15.10 | 3.42 | -5.62 | 12.45 |
| 11 | Brain cancer | 4.21 | -7.05 | 15.47 | -2.21 | -13.33 | 8.90 |
| 12 | Leukaemia / Lymphoma | 3.67 | 0.02 | 7.33 | -0.48 | -4.08 | 3.13 |
| 13 | **Blood diseases** | 5.07 | 3.33 | 6.82 | 6.69 | 4.98 | 8.40 |
| 14 | Anaemia | 4.78 | 2.89 | 6.66 | 6.97 | 5.11 | 8.82 |
| 15 | **Endocrine diseases** | 2.52 | 1.52 | 3.52 | 4.32 | 3.33 | 5.30 |
| 16 | Diabetes mellitus | 6.75 | 4.20 | 9.29 | 11.42 | 8.95 | 13.89 |
| 17 | **Mental and behavioural disorders** | 3.93 | 2.61 | 5.26 | 5.45 | 4.16 | 6.75 |
| 18 | Dementia | 1.20 | -3.30 | 5.70 | 7.43 | 2.96 | 11.89 |
| 19 | Substance use disorders | 4.17 | 2.17 | 6.17 | 6.60 | 4.65 | 8.55 |
| 20 | Psychotic disorders | 7.46 | -3.04 | 17.96 | 14.05 | 3.77 | 24.33 |
| 21 | Mood disorders | 5.15 | 2.92 | 7.38 | 5.39 | 3.20 | 7.58 |
| 22 | Neurotic disorders | 4.37 | 2.09 | 6.64 | 4.86 | 2.62 | 7.11 |
| 23 | **Neurological disorders** | 0.94 | -0.56 | 2.44 | 2.54 | 1.07 | 4.01 |
| 24 | Parkinson disease | 2.52 | -4.30 | 9.35 | 3.15 | -3.58 | 9.88 |
| 25 | Multiple sclerosis | 9.07 | -0.19 | 18.32 | 6.21 | -2.87 | 15.29 |
| 26 | Epilepsy | 3.46 | -2.06 | 8.99 | 7.64 | 2.25 | 13.03 |
| 27 | Headaches | 1.30 | -2.68 | 5.29 | 0.66 | -3.29 | 4.61 |
| 28 | Transient ischaemic attack | 0.30 | -5.70 | 6.29 | 4.03 | -1.85 | 9.91 |
| 29 | Sleep disorders | 2.44 | -2.76 | 7.65 | 0.45 | -4.65 | 5.56 |
| 30 | **Eye diseases** | 1.58 | 0.33 | 2.84 | 2.17 | 0.93 | 3.42 |
| 31 | **Ear diseases** | 0.53 | -6.23 | 7.29 | 2.96 | -3.65 | 9.56 |
| 32 | **Circulatory diseases** | 1.58 | 0.80 | 2.37 | 2.16 | 1.39 | 2.93 |
| 33 | Hypertension | 2.22 | 1.19 | 3.25 | 4.80 | 3.79 | 5.81 |
| 34 | Ischaemic heart disease | 1.57 | -0.37 | 3.50 | 5.14 | 3.25 | 7.02 |
| 35 | *Angina pectoris* | 0.68 | -2.15 | 3.51 | 6.70 | 3.96 | 9.44 |
| 36 | *Myocardial infarction* | 0.19 | -3.54 | 3.91 | 6.41 | 2.81 | 10.00 |
| 37 | Pulmonary embolism | 6.52 | 1.97 | 11.06 | -0.58 | -5.13 | 3.98 |
| 38 | Arrhythmias | 4.25 | 2.20 | 6.30 | -3.74 | -5.78 | -1.69 |
| 39 | Heart failure | 5.99 | 2.50 | 9.48 | 4.55 | 1.09 | 8.01 |
| 40 | Cerebrovascular diseases | 4.56 | 1.72 | 7.41 | 7.33 | 4.53 | 10.13 |
| 41 | Stroke | 3.03 | -0.67 | 6.73 | 4.34 | 0.69 | 7.98 |
| 42 | *Intracerebral haemorrhage* | 2.87 | -3.59 | 9.34 | 3.49 | -2.88 | 9.86 |
| 43 | *Cerebral infarction* | 3.36 | -1.00 | 7.72 | 4.80 | 0.50 | 9.09 |
| 44 | Arteriosclerosis | 5.73 | -1.91 | 13.37 | 13.10 | 5.65 | 20.55 |
| 45 | Deep vein thrombosis | 4.39 | -0.36 | 9.14 | -1.83 | -6.55 | 2.90 |
| 46 | **Respiratory diseases** | 5.62 | 4.44 | 6.79 | 6.41 | 5.26 | 7.57 |
| 47 | Respiratory tract infections | 8.97 | 6.54 | 11.39 | 10.11 | 7.72 | 12.50 |
| 48 | Chronic obstructive pulmonary disease | 11.11 | 8.32 | 13.90 | 14.70 | 11.94 | 17.46 |
| 49 | Asthma | 5.93 | 3.92 | 7.94 | 6.86 | 4.89 | 8.82 |
| 50 | **Digestive system diseases** | 2.55 | 1.81 | 3.30 | 2.89 | 2.16 | 3.62 |
| 51 | Appendicitis | 0.17 | -6.84 | 7.18 | -4.21 | -11.04 | 2.61 |
| 52 | Inflammatory bowel disease | 4.34 | 1.61 | 7.07 | 5.66 | 2.99 | 8.33 |
| 53 | Liver diseases | 6.77 | 3.21 | 10.33 | 9.55 | 6.07 | 13.03 |
| 54 | Alcohol-related liver diseases | 19.25 | 7.64 | 30.85 | 30.04 | 18.99 | 41.09 |
| 55 | Pancreatitis | 10.10 | 2.30 | 17.90 | 11.02 | 3.36 | 18.68 |
| 56 | **Skin diseases** | 2.74 | 1.14 | 4.35 | 1.61 | 0.03 | 3.19 |
| 57 | Eczema and skin infections | 3.61 | 0.81 | 6.40 | 4.10 | 1.36 | 6.84 |

| | | | | | | |
|---|---|---|---|---|---|---|
| 58 **Musculoskeletal diseases** | 0.00 | -0.86 | 0.85 | 0.11 | -0.72 | 0.95 |
| 59 Rheumatoid arthritis | 2.68 | 1.23 | 4.13 | 4.12 | 2.68 | 5.56 |
| 60 Gout | 2.19 | -2.81 | 7.19 | 3.33 | -1.51 | 8.17 |
| 61 Osteoarthritis | -2.30 | -3.61 | -0.98 | -2.80 | -4.10 | -1.50 |
| 62 Sciatica | -0.81 | -4.01 | 2.39 | -2.21 | -5.36 | 0.94 |
| 63 Backpain | 1.10 | -1.34 | 3.53 | 2.00 | -0.40 | 4.39 |
| 64 Soft tissue diseases | -0.13 | -1.86 | 1.61 | 0.04 | -1.66 | 1.74 |
| 65 **Genitourinary diseases** | 3.60 | 2.48 | 4.72 | 3.11 | 2.02 | 4.21 |
| 66 Renal failure | 4.18 | 1.98 | 6.37 | 4.59 | 2.42 | 6.76 |
| 67 **Circulatory and respiratory symptoms** | 2.60 | 1.16 | 4.04 | 2.73 | 1.31 | 4.14 |
| 68 **Digestive and abdominal symptoms** | 2.58 | 1.32 | 3.85 | 3.36 | 2.12 | 4.59 |
| 69 **Injury** | 3.24 | 1.63 | 4.85 | 2.77 | 1.19 | 4.34 |
| 70 **Poisoning** | 2.65 | -3.91 | 9.20 | 6.17 | -0.22 | 12.55 |
| 71 **Road accidents** | -2.58 | -7.55 | 2.38 | -4.78 | -9.52 | -0.04 |
| 72 **Fall** | 5.15 | 3.17 | 7.13 | 4.65 | 2.69 | 6.61 |
| 73 **Self-harm** | 0.12 | -8.31 | 8.54 | 4.61 | -3.52 | 12.75 |

Supplement eTable 8. Associations of obesity with outcomes with minimal adjustment

| | HR for obesity vs normal weight defined using | | |
|---|---|---|---|
| **Outcome** | **BMI** | **WC** | **WHtR** |
| **UK Biobank** | | | |
| All-cause mortality | 1.65 (1.60-1.70) | 1.66 (1.61-1.71) | 1.66 (1.61-1.71) |
| Simple multimorbidity (2$^{nd}$ disease) | 2.99 (2.95-3.04) | 3.00 (2.95-3.04) | 3.00 (2.95-3.05) |
| Third disease | 3.72 (3.65-3.80) | 3.73 (3.66-3.81) | 3.74 (3.66-3.81) |
| Complex multimorbidity (4$^{th}$ disease) | 4.58 (4.46-4.70) | 4.60 (4.48-4.72) | 4.60 (4.48-4.72) |
| **Whitehall** | | | |
| All-cause mortality | 1.81 (1.59-2.04) | 1.82 (1.61-2.06) | 1.92 (1.70-2.17) |
| Simple multimorbidity (2$^{nd}$ disease) | 2.50 (2.27-2.75) | 2.61 (2.37-2.87) | 2.60 (2.36-2.86) |
| Third disease | 2.68 (2.39-3.00) | 2.78 (2.49-3.11) | 2.77 (2.47-3.10) |
| Complex multimorbidity (4$^{th}$ disease) | 3.36 (2.95-3.83) | 3.38 (2.97-3.85) | 3.42 (3.00-3.90) |

Numbers presented are HR (95% CI) compared with the normal weight group and its WC/WHtR equivalent.
Adjusted for age, sex, and ethnicity.

Supplement eTable 9. Difference in population attributable fractions between obesity defined using different measures with minimal adjustment

| | Obesity (BMI ≥ 30) | | | | | | Overweight (BMI ≥ 25 & < 30) | | | | | |
|---|---|---|---|---|---|---|---|---|---|---|---|---|
| | $PAF_{WC}$ - $PAF_{BMI}$ | | | $PAF_{WHtR}$ - $PAF_{BMI}$ | | | $PAF_{WC}$ - $PAF_{BMI}$ | | | PAFWHtR - PAFBMI | | |
| **Outcome** | **%** | **Lower CI** | **Upper CI** | **%** | **Lower CI** | **Upper CI** | **%** | **Lower CI** | **Upper CI** | | **Lower CI** | **Upper CI** |
| **UK Biobank** | | | | | | | | | | | | |
| All-cause mortality | 0.21 | -1.79 | 2.21 | 0.22 | -1.78 | 2.22 | 7.80 | 5.71 | 9.88 | 10.62 | 8.58 | 12.66 |
| Simple multimorbidity (2nd disease) | 0.07 | -0.73 | 0.86 | 0.07 | -0.72 | 0.87 | 2.51 | 1.66 | 3.37 | 3.12 | 2.28 | 3.96 |
| Third disease | 0.13 | -0.82 | 1.08 | 0.14 | -0.81 | 1.10 | 4.68 | 3.51 | 5.86 | 5.67 | 4.52 | 6.83 |
| Complex multimorbidity (4th disease) | 0.18 | -0.94 | 1.29 | 0.19 | -0.92 | 1.31 | 6.31 | 4.71 | 7.90 | 7.78 | 6.21 | 9.36 |
| **Whitehall** | | | | | | | | | | | | |
| All-cause mortality | -0.19 | -1.41 | 0.91 | 0.75 | -0.56 | 2.03 | 4.27 | 1.49 | 7.43 | 2.67 | -0.23 | 5.60 |
| Simple multimorbidity (2nd disease) | 0.47 | -0.90 | 1.94 | 0.60 | -0.90 | 2.00 | 1.86 | -0.33 | 3.72 | 0.72 | -1.32 | 2.86 |
| Third disease | 0.34 | -1.34 | 1.87 | 0.38 | -1.19 | 1.88 | 1.92 | -0.93 | 4.46 | 1.49 | -1.17 | 3.99 |
| Complex multimorbidity (4th disease) | -0.23 | -2.45 | 1.40 | 0.02 | -2.01 | 1.81 | 1.81 | -1.74 | 4.73 | 2.02 | -1.11 | 5.24 |

PAF assumes the HR estimated to be causal which may not be true.

HRs were adjusted for age, sex, and ethnicity.

**eFigure 1 Associations of adiposity markers with infections and cancer outcomes**

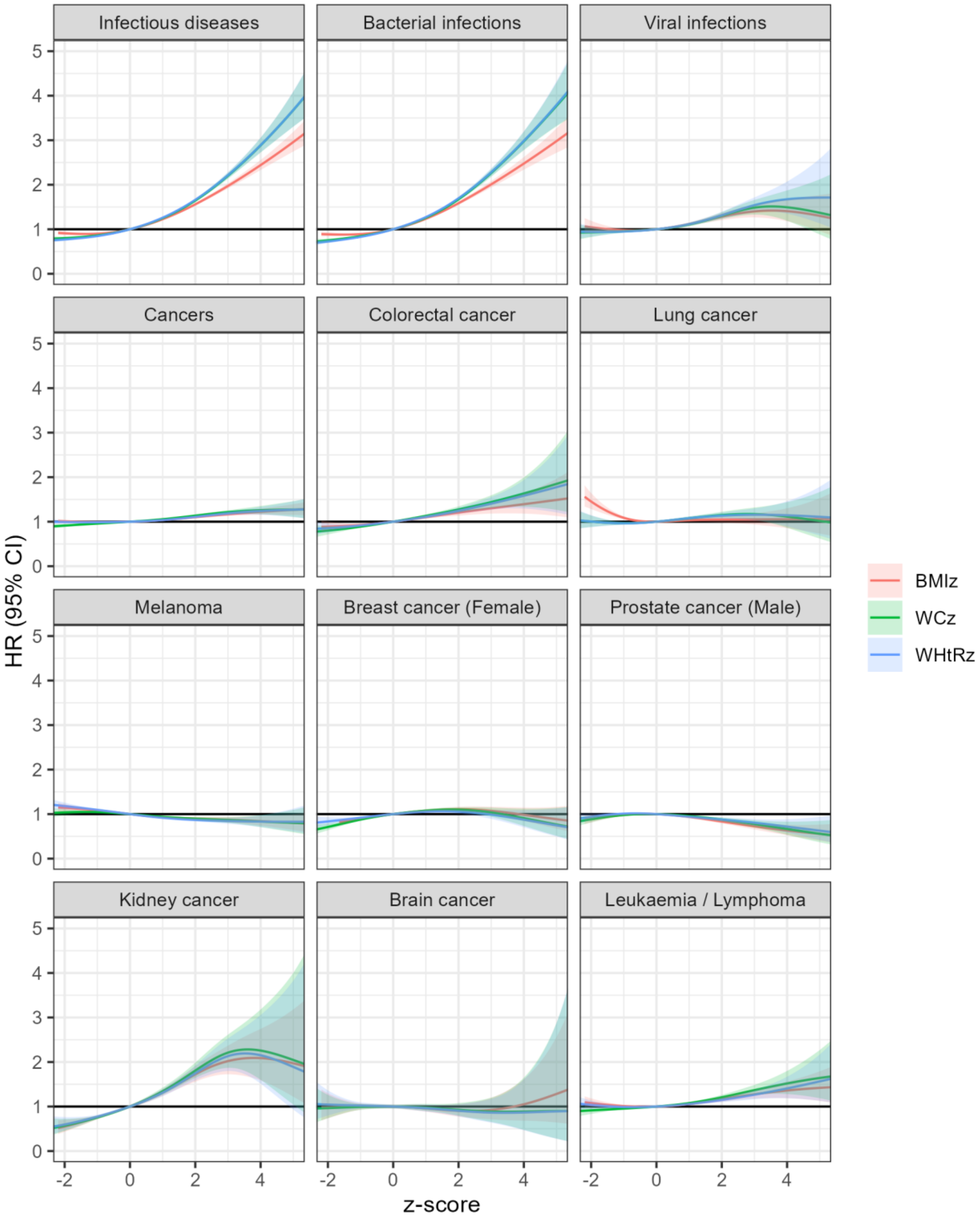

**eFigure 2 Associations of adiposity markers with blood and endocrine and mental health outcomes**

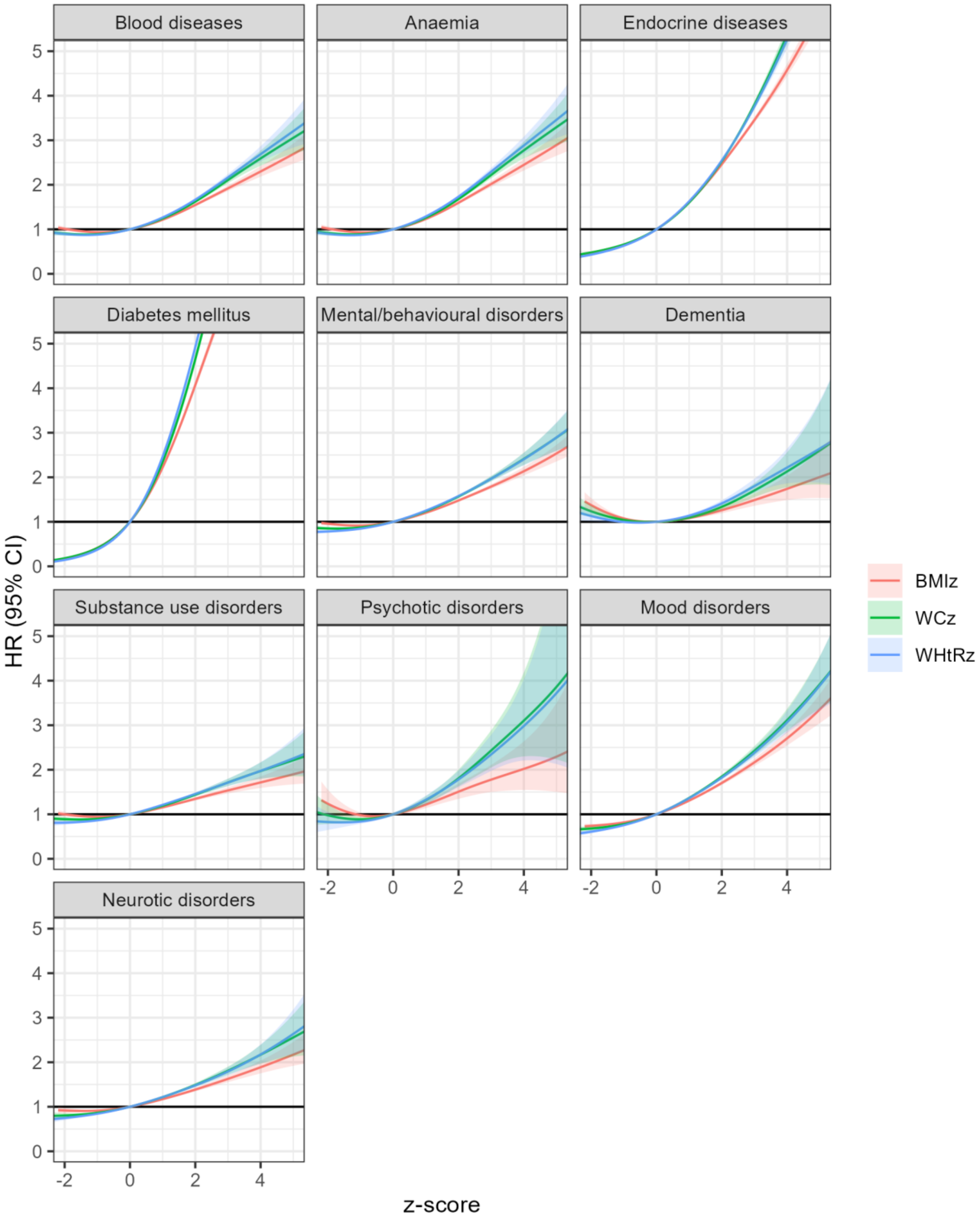

**eFigure 3 Associations of adiposity markers with neuro and eye and ear outcomes**

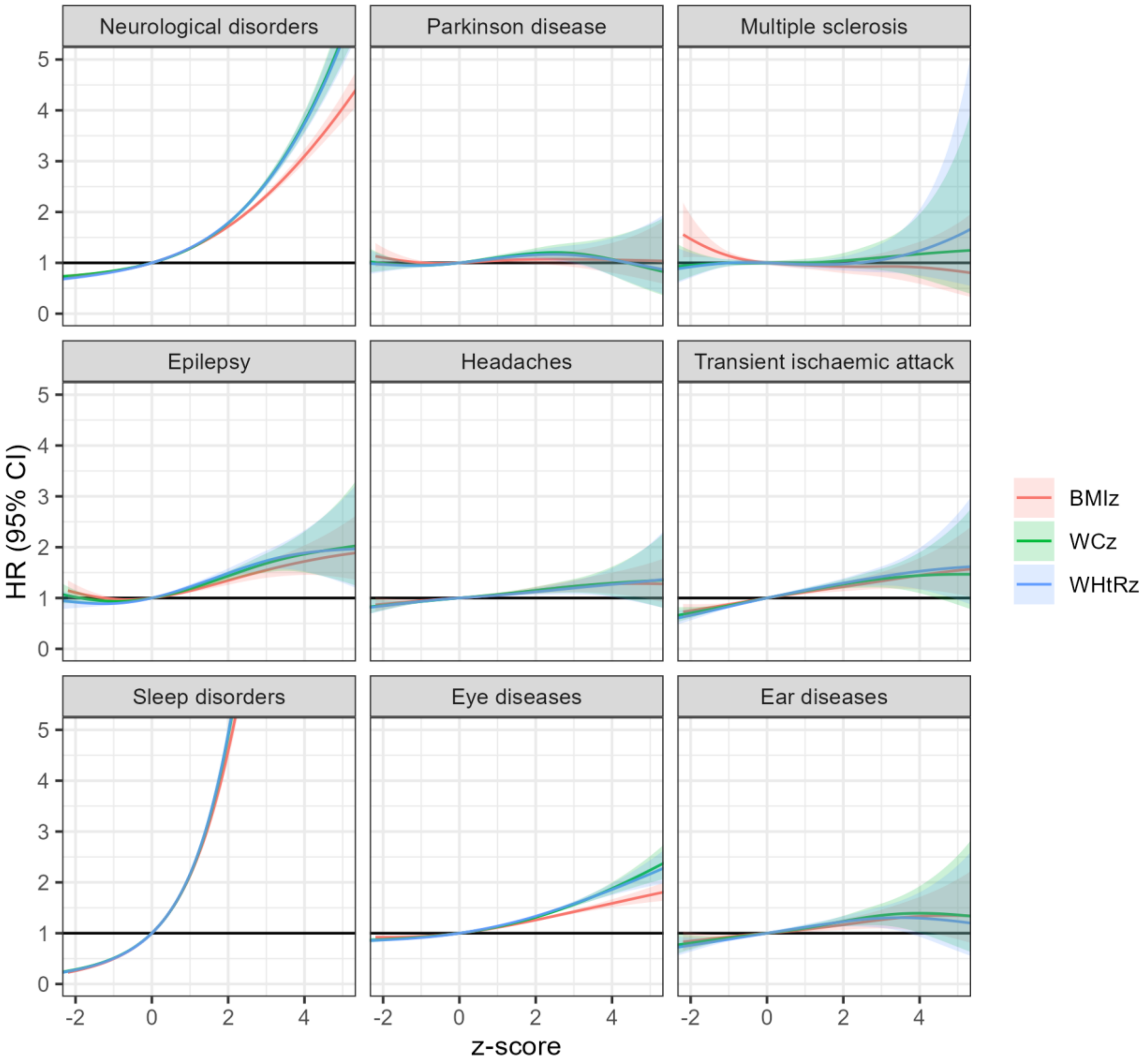

**eFigure 4 Associations of adiposity markers with circulatory outcomes**

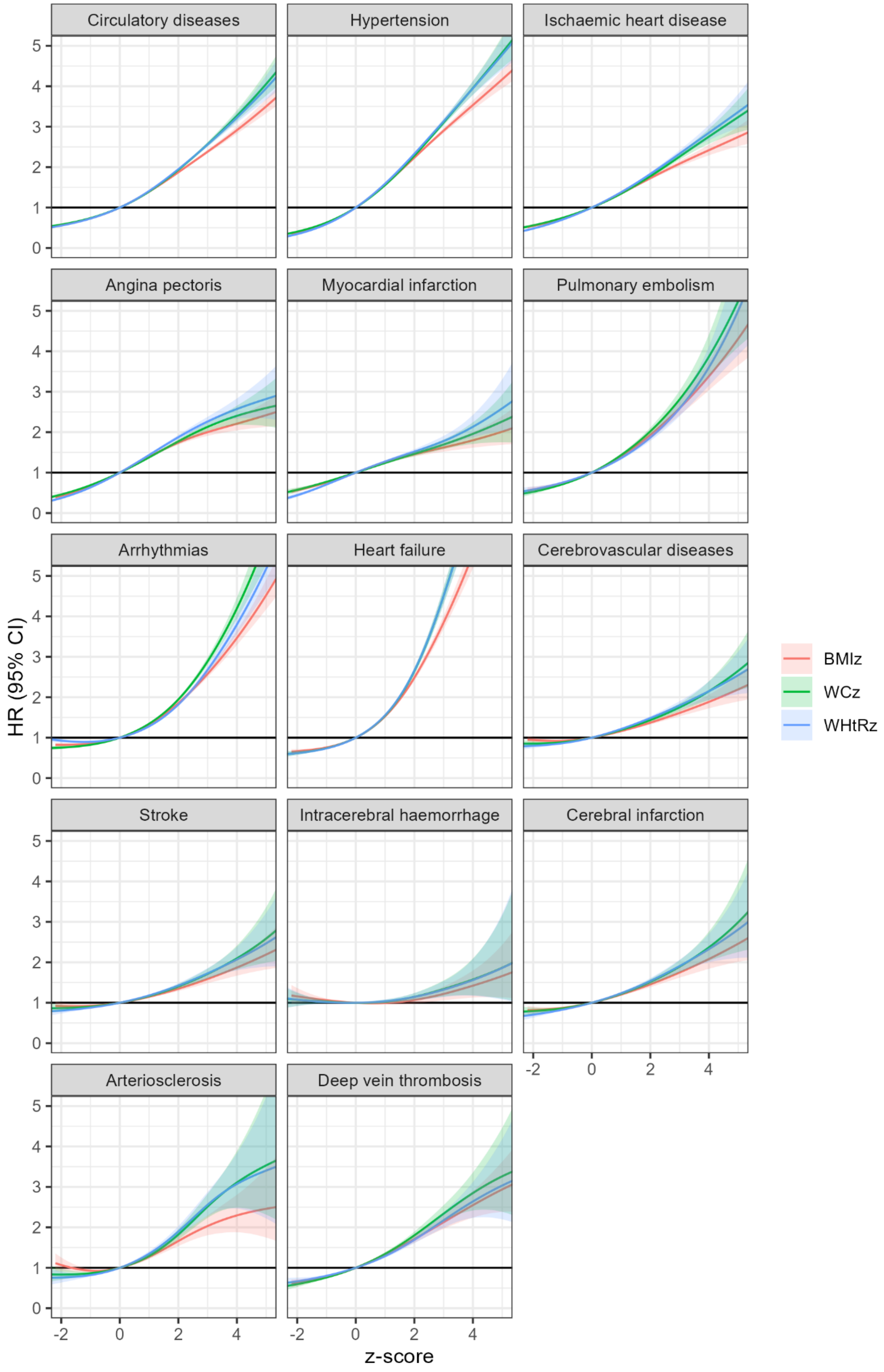

**eFigure 5 Associations of adiposity markers with respiratory and digestive outcomes**

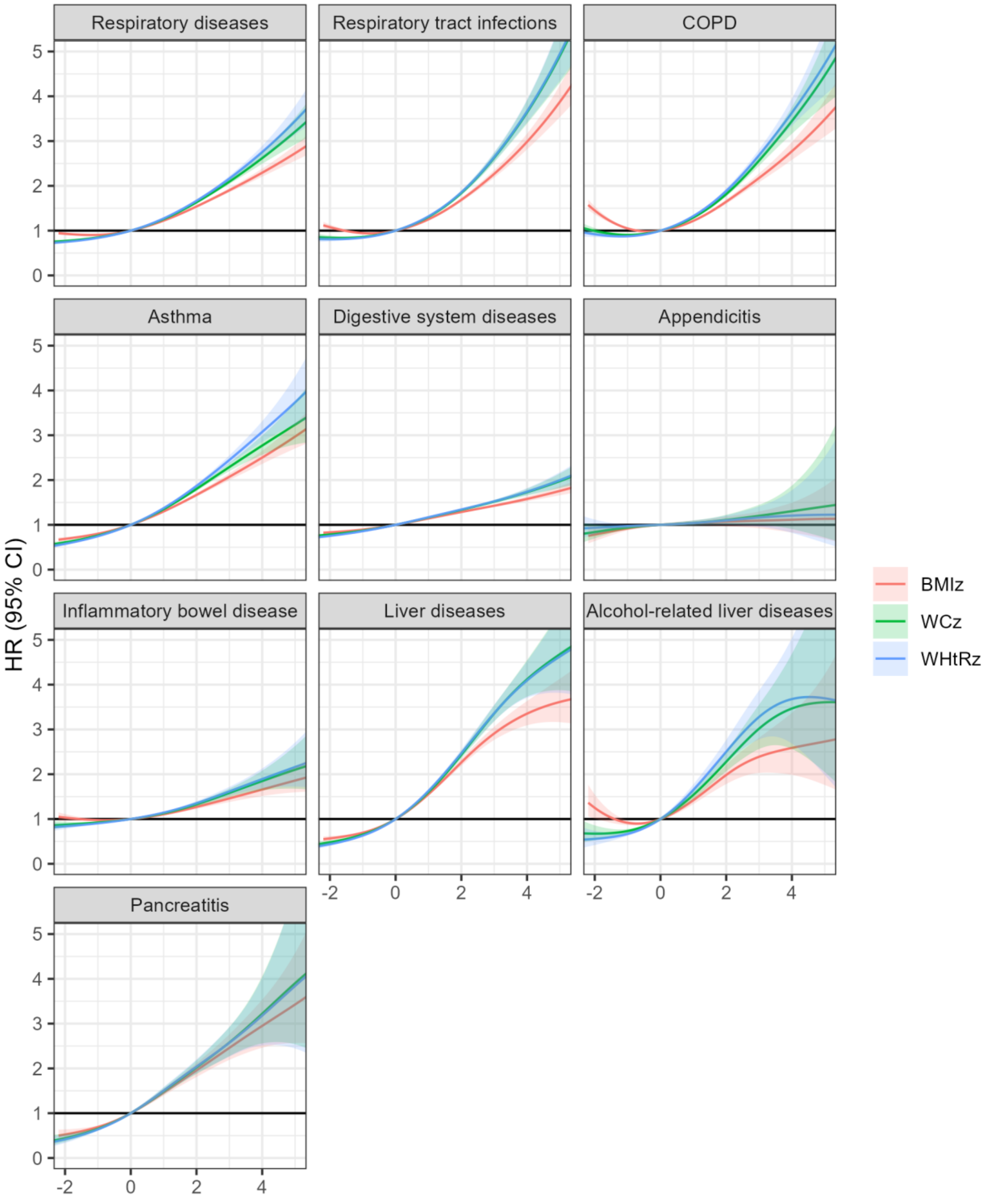

**eFigure 6 Associations of adiposity markers with skin and musculoskeletal outcomes**

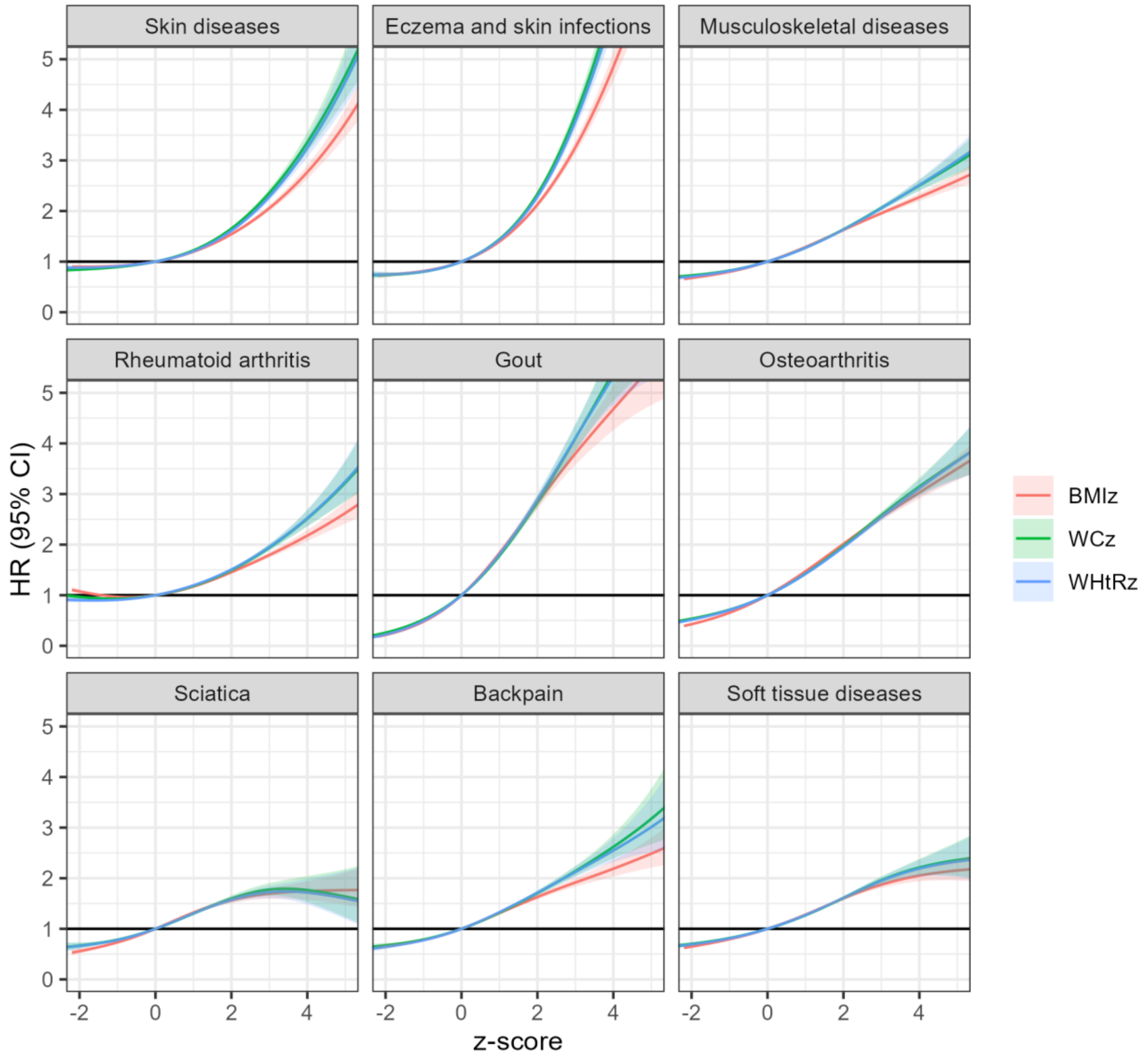

**eFigure 7 Associations of adiposity markers with genitourinary and miscellaneous outcomes**

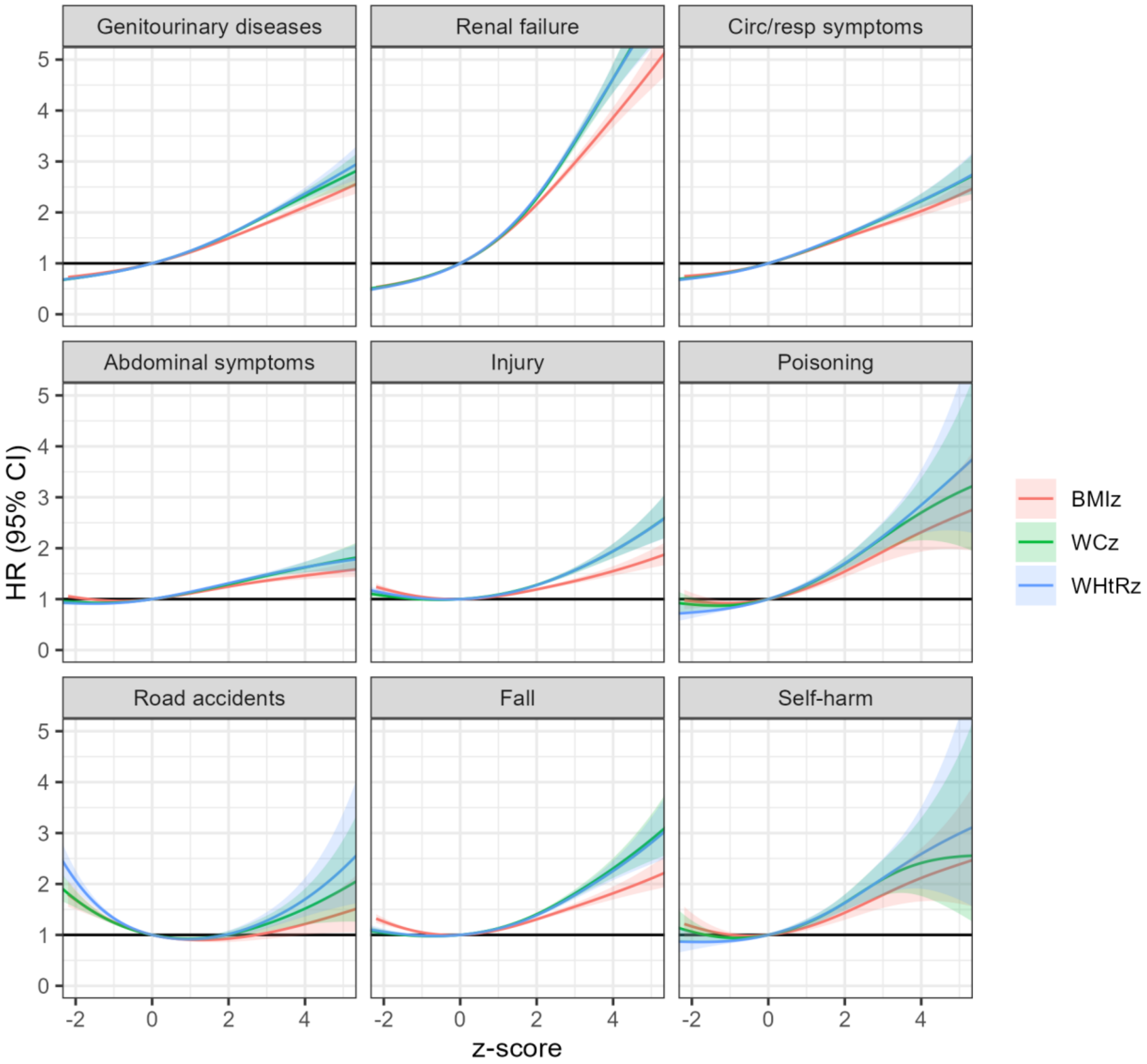
